# Large-scale admixture mapping in the *All of Us Research Program* improves the characterization of cross-population phenotypic differences

**DOI:** 10.1101/2025.04.02.25325115

**Authors:** Ravi Mandla, Zhuozheng Shi, Kangcheng Hou, Ying Wang, Georgia Mies, Alan J. Aw, Sinead Cullina, Penn Medicine BioBank, Eimear Kenny, Elizabeth Atkinson, Alicia R. Martin, Bogdan Pasaniuc

**Affiliations:** Graduate Program in Genomics and Computational Biology, University of Pennsylvania, Philadelphia, PA, USA; Department of Epidemiology, Harvard T.H. Chan School of Public Health, Boston, MA, USA; Analytic and Translational Genetics Unit, Massachusetts General Hospital, Boston, MA, USA; Stanley Center for Psychiatric Research, Broad Institute of MIT and Harvard, Cambridge, MA, USA; Program in Medical and Population Genetics, Broad Institute of MIT and Harvard, Cambridge, MA, USA; Department of Genetics, Perelman School of Medicine, University of Pennsylvania, Philadelphia, PA, USA; Institute for Genomic Health, Icahn School of Medicine at Mount Sinai, New York, NY, USA; Department of Genetics and Genomic Sciences, Icahn School of Medicine at Mount Sinai, New York, NY, USA; Department of Molecular and Human Genetics, Baylor College of Medicine, Houston, TX, USA

**Author notes:** A full list of consortium members is provided in the supplemental information (Penn Medicine BioBank). These authors contributed equally to this work (Ravi Mandla, Zhuozheng Shi).

## Abstract

Admixed individuals have largely been understudied in medical research due to their complex genetic ancestries. However, the consideration of admixture can help identify ancestry-enriched genetic associations, delineating some of the genetic underpinnings of cross-population phenotypic variation.

To this end, we performed local ancestry inference within the *All of Us Research Program* to identify individuals with recent admixture between African (AFR) and European (EUR) populations (N=48,921). We identified evidence of local AFR ancestry enrichment at the HLA locus, suggestive of putative selection since admixture. Furthermore, we performed the largest admixture mapping (ADM) efforts in AFR-EUR Admixed individuals for 22 traits, identifying 71 associations between inferred local AFR ancestries and a trait. Variants from published GWAS could only account for 18 (25%) of the ADM associations, highlighting novel loci where ancestral haplotypes explained some phenotypic variation. Previous studies likely have not identified these loci due to the low availability of high-powered GWAS in populations genetically similar to AFR. One such loci was 9q21.33, associated with 1.4-fold risk of end-stage kidney disease (ESKD) for carriers of inferred local AFR ancestries at the region. This locus contains the gene *SLC28A3*, which has previously been linked to kidney function but has never been associated with cross-population ESKD prevalence differences.

Together, our results expand upon the existing literature on phenotypic differences between populations, highlighting loci where genetic ancestries play a critical role in the genetic architecture of disease.

## Introduction

Genome wide association studies (GWAS) have successfully identified thousands of genetic loci associated with a range of diseases and related quantitative traits over the past 15 years^1^. GWAS results are increasingly being leveraged into potential clinical insights, including the identification of novel drug targets^2,3^ and improved disease risk prediction using polygenic scores (PGS)^4,5^. Historically, many of these studies were conducted within populations genetically similar to European individuals^6^. However, growing recognition of the importance of diversifying genetics research has spurred multiple global initiatives to include more underrepresented population groups within genetic studies^7–15^. Such efforts have led to improvements in PGS portability^16,17^, fine-mapping^18,19^, and the identification of GWAS signals with cross-ancestry heterogeneity^7,20^.

Despite broader inclusion of these populations, genetic analyses largely continue to exclude recently admixed individuals, defined here as individuals which underwent a mixture of multiple diverged continental populations within the last 20 generations. The genomes of these individuals contain many segments from these different ancestral populations with differing linkage-disequilibrium (LD) and allele frequency (AF) patterns, referred to as local ancestry, which can complicate population structure adjustments in GWAS. Inclusion of admixed individuals can also make downstream uses of GWAS, such as fine-mapping and colocalization methods, more difficult as they require reference panels from pre-defined ancestral populations^21–23^. However, as the number of admixed individuals continues to increase, improving their representation in genetic analyses is crucial for translating genetic insights into healthcare settings.

The unique genomes of admixed individuals can also shed light on the genetic architecture of traits. For example, previous work has explored how GWAS from different populations can be meta-analyzed together to identify “heterogeneous” signals^24^. However, non-genetic differences between the populations such as environmental and ascertainment bias also confound such insights, muddying the interpretation of results. These biases can be attenuated when studying admixed populations, which tend to share similar environmental and ascertainment contexts but contain different genetic segments with “local ancestry” from multiple populations. This allows for more robust associations of cross-population genetic variations with traits of interest^25^. One such method, called admixture mapping (ADM), is well-powered for identifying trait-associated loci with different allele frequencies between ancestral populations^26,27^, and has previously identified several important genes relevant to disease prevalence differences such as the *APOL1* locus in end stage kidney disease (ESKD)^28,29^. Furthermore, deviations in local ancestry proportions from genome-wide averages may indicate regions that have undergone selection since admixture^30^. Historically, however, ADM has had several limitations. First, the small sample size of admixed individuals in most genetic datasets have limited the statistical power for ADM^31^. Second, most ADM initiatives have been performed in specific disease cohorts, preventing robust multi-trait comparisons that could guide ideal use cases of ADM. Finally, despite increases in the diversity of biobanks, the lack of inferred local ancestry data prevents larger, standardized genetic analyses within admixed individuals.

Here, we describe for the first time inferred local ancestry data within the *All of Us Research Program* (AoU), a diverse US-based biobank with linked genetic, electronic health record (EHR) data, and survey information^13,32^. Leveraging this local ancestry data, we identified 48,921 genetically-inferred two-way African-European admixed individuals (AFR-EUR Admixed) and found evidence of selection since admixture around the HLA locus. We then performed the largest ADM initiative in AFR-EUR Admixed individuals with 22 binary and quantitative traits, discovering over 50 genetic loci associated with at least one phenotype. Many of our associations were independent of an established GWAS signal, including loci which have never been associated with specific disease-relevant traits before. Together, these results highlight the power of including admixed individuals in genetic studies to uncover novel biological mechanisms that underly cross-population trait variations.

## Results

### Characterization of admixture within The All of Us Research Program

To assess levels of admixture within the US, we pulled common variants from short read whole genome sequencing (srWGS) data for 245,394 AoU participants, then performed global and local ancestry inference using four super population labels from the 1000 Genomes Project (1000G; African: AFR; European: EUR; East Asian: EAS; South Asian: SAS) and 1000G samples from Lima, Peru, which we refer to as Native American (NAT; **Methods**). Using inferred global ancestry proportions, we subset the data to unrelated AFR-EUR Admixed individuals (N=48,921; **Fig. 1A**; **Fig. 1B**; **Methods**). We observed an 85% overlap between the AFR-EUR Admixed individuals and participants whose self-identified race and ethnicity (SIRE) is non-Hispanic Black or African American (**Extended Data Fig. 1**; **Supplementary Table 1**), suggestive of high similarity between genetically inferred admixed individuals and self-reported race/ethnicity. We note AFR-EUR Admixed individuals were also closer to EUR individuals compared to SIRE non-Hispanic Black or African American individuals within principal component (PC) space (**Extended Data Fig. 2**), indicating a larger inclusion of admixed individuals closer to European ancestry compared to SIRE non-Hispanic Black or African American subsets.

**Fig. 1.**
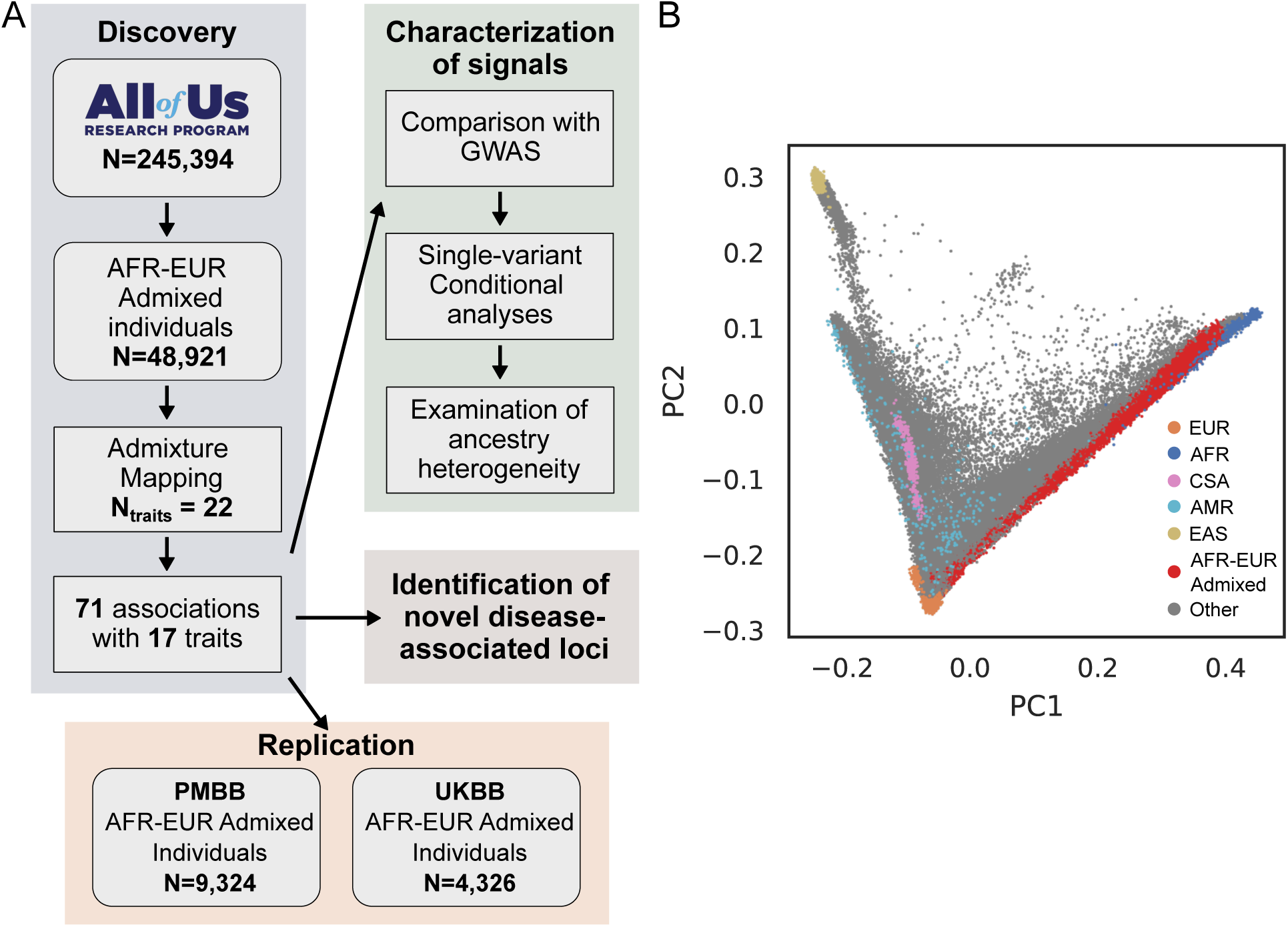
Overview of admixture mapping in All of Us. (A) Workflow of the study design for performing admixture mapping in All of Us (AoU). (B) AoU participants plotted in PC space, along with reference populations from 1000G. AoU participants who are not classified as AFR-EUR Admixed are colored grey. PMBB: Penn Medicine Biobank. UKBB: UK Biobank

Previous studies have explored selection within admixed individuals^28,30,33^, where genomic loci with local ancestry frequencies significantly differing from the average over the whole genome could indicate regions under putative selection since admixture. We tested for evidence of selection in AFR-EUR Admixed individuals within AoU, a dataset almost twice as large as those previously used for similar analyses^33^ (**Methods**). With both the same QC steps and p-value cutoff previously described^33^, we found the inferred European ancestry was significantly depleted in the HLA locus relative to the rest of the genome with an estimated lower bound selection coefficient of 0.037 (**Fig. 2**; **Methods**). We observed similar results when using SIRE non-Hispanic Black or African American individuals (**Extended Data Fig. 3**). As replication, we also tested if such variations exist in other AFR-EUR Admixed populations. Indeed, within the Penn Medicine Biobank (PMBB; N=9,324), we still observed significant decreases in inferred local EUR ancestries with an estimated lower bound selection coefficient of 0.015, even after masking the HLA locus (**Extended Data Fig. 4**; **Methods**). To determine if there were any phenotypic consequences of this candidate selection, we tested for associations between inferred European ancestry at the HLA locus and phecodes and immune-relevant labs within AoU (**Methods**). After adjusting for multiple-hypothesis testing, we did not observe any significant associations (**Supplementary Table 2**; **Supplementary Table 3**; **Methods**).

**Fig. 2.**
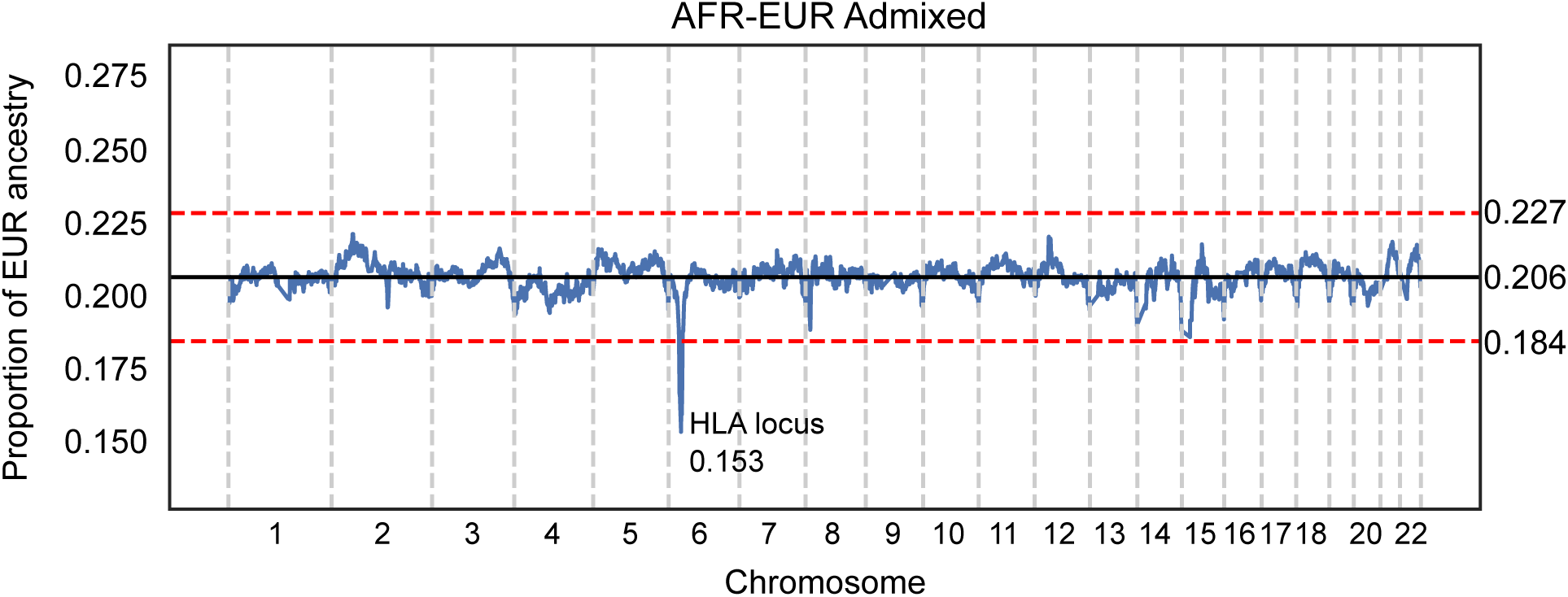
Putative evidence of selection since admixture. Proportion of local EUR ancestry across the genome in unrelated genetically-inferred AFR-EUR Admixed participants. The solid black line represents the genome-wide average proportion of local EUR ancestry, and the red dashed lines represent the genome-wide significant P<0.05 cutoff of 4.42 SD away from the mean.

### Identification of genomic loci associated with trait differences across populations

Evaluating local ancestry associations with various phenotypes can highlight novel trait-associated loci and shed light on cross-ancestry genetic heterogeneity. To this end, we performed ADM within AFR-EUR Admixed individuals for 22 traits (7 binary and 15 quantitative; **Methods**). ADM identified a total of 59 independent loci significantly associated with 17 (77.3%) of the traits tested (P<5×10^−5^), for a total of 71 different loci-to-trait associations (**Fig. 3A**, **3B**; **Supplementary Table 4**; **Methods**). Among these hits included well-characterized admixture associations, such as that between the Duffy locus and neutrophil count^34^ and the *APOL1* locus and ESKD^28,29,35^ (**Supplementary Table 4**). As QC, we tested whether errors in local ancestry inference due to low-complexity regions^36^ could explain some of these associations, and found minimal overlap between our associated loci and low-complexity regions (**Supplementary Table 5**; **Methods**). Twelve (20.4%) of the loci were associated with multiple traits, suggestive of shared cross-population genetic architecture for related traits. We tested these associations for evidence of pairwise colocalization, defined as having a posterior probability of sharing a causal signal greater than 0.8. In total, 11 (91.7%) of the multi-trait associated loci demonstrated evidence of colocalization, which is expected given the high correlation between many of the traits (i.e. hip circumference and BMI; **Supplementary Table 5**; **Methods**). More broadly, we found evidence of genetic correlation between related traits across the 59 loci, including HbA1c and type 2 diabetes (Pearson R=0.6; P=5.4×10^−8^; **Extended Data Fig. 5**; **Methods**), indicating pleiotropy of admixture signals between traits.

**Fig 3.**
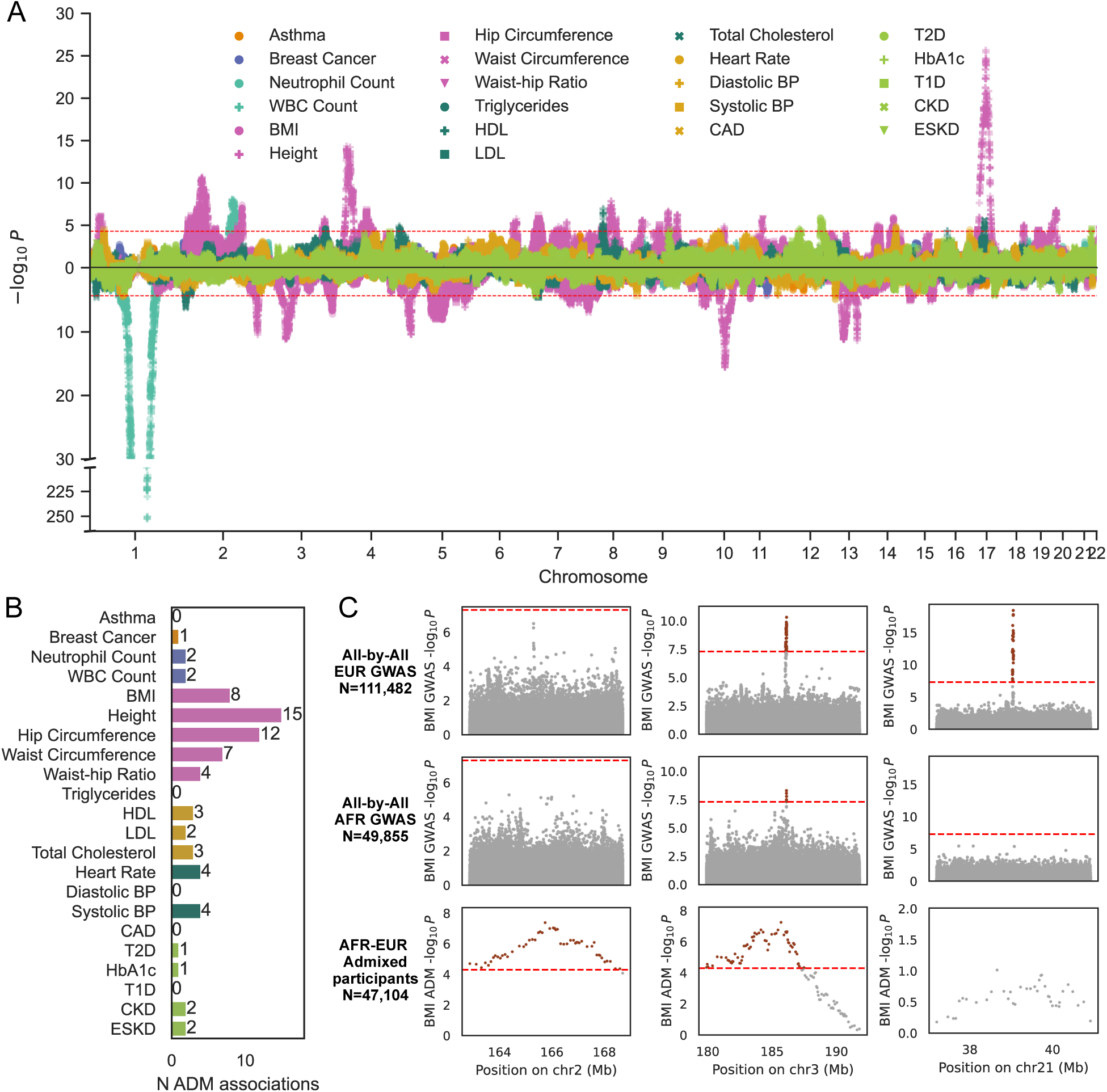
Summary of admixture mapping results. (A) Miami plot of admixture mapping (ADM) results across all traits, with effect directions relative to AFR local ancestry haplotypes. The dashed lines represent the genome-wide significant cutoff (P<5×10^−5^). Traits are colored based on phenotype category, including orange for respiratory, dark blue for cancer, light blue for immune, pink for anthropometric, dark green for lipid, yellow for circulatory, and light green for metabolic/endocrine traits. The shape of the dots indicates the unique traits within each phenotype category. (B) Bar plot of the number of genome-wide significant ADM peaks per trait. (C) Example loci detected using GWAS and/or ADM for BMI. Dashed red lines represent the genome-wide significance cutoffs for GWAS (P<5×10^−8^) or ADM (P<5×10^−5^). Red dots represent associations which pass the genome-wide significance cutoff.

As an example, ADM in AoU identified 8 loci where inferred local African ancestry is significantly associated with BMI. When comparing these results to available All by All AoU GWAS in participants AoU previously denoted as genetically similar to European and African reference populations^32^, we found 6 loci with ADM significance only (**Fig. 3C, left**), 2 loci identified also with GWAS (**Fig. 3C, center**), and additional loci ADM fails to identify but are found in a GWAS, even if the loci is differentially significant between subgroups (**Fig. 3C, right**) when using the traditional genome-wide significance threshold for GWAS (P<5×10^−8^). These latter examples may be due to several factors including limited power of current available sample sizes for ADM, loci without extensive admixture between African and European populations, or causal variants with similar frequencies on AFR and EUR haplotypes.

For replication, we conducted ADM on 20 of the 22 previously defined traits (4 binary and 16 quantitative) in 4,326 AFR-EUR Admixed individuals from the UK Biobank (UKBB) (**Supplementary Table 6**; **Extended Data Fig. 6**; **Methods**) and on 19 of the 22 traits (7 binary and 12 quantitative) in 9,324 previously-defined AFR-EUR Admixed individuals from the Penn Medicine Biobank (**Supplementary Table 8**; **Methods**)^37^. We found that 2 of the associations replicated in UKBB and 6 in PMBB (**Supplementary Table 7**; **Supplementary Table 9**; **Extended Data Fig. 7A**; **Methods**). This included associations between white blood cell and neutrophil count and the Duffy locus, as well as various associations with height and BMI. While the number of replications was low, effect size estimates were strongly correlated and had consistent effect directions between AoU and UKBB (Pearson R: 0.5; P=6.5×10^−5^) and between AoU and PMBB (Pearson R: 0.5; P=2.5×10^−4^), especially for nominal associations (P<0.05; UKBB: Pearson R=0.7, P=3.5×10^−3^; PMBB: Pearson R=0.6, P=3.5×10^−4^), suggesting shared signals but limited power in UKBB and PMBB (**Extended Data Fig. 7B, 7C**).

### Genetic architecture underlying ADM associations

Previous studies have also demonstrated that ADM is most powered to detect causal signals with differing allele frequencies between ancestries^27^. To characterize the underlying signals we capture with ADM, we re-ran ADM conditioning on variants reported to be significantly associated with the same trait in the GWAS catalog^38^ (**Supplementary Table 10**; **Methods**). Of the 71 ADM associations, 69 (97%) were near a published GWAS signal and 18 (25.4%) decreased in significance after conditional analysis (*P*>1×10^−4^), suggesting the conditioned variant may partially explain the ADM signal (**Fig. 4A**). We found these conditioned variants had a significantly larger absolute difference between the alternate allele frequencies (altAF) in AFR and EUR populations when compared to variants which did not decrease ADM significance after conditioning (Mann-Whitney U *P*=1.0×10^−4^; **Fig. 4B**). Variants which did not condition an ADM association were also more common in EUR populations compared to AFR populations, suggesting conditional analyses may be underpowered for some of these variants since average EUR global ancestry proportions in the AoU AFR-EUR Admixed populations were smaller than the average AFR global ancestry proportions (**Fig. 2**). Variants which conditioned an ADM association also had a significantly larger absolute log fold change between the altAFs of AFR and EUR populations (Mann-Whitney U *P*=3.2×10^−4^; **Extended Data Fig. 8A**). When comparing the effect sizes of these variants from their associations with the traits in AoU participants genetically similar to AFR (AFR-like) and EUR (EUR-like) populations (**Methods**), we also found significantly larger effect size differences for variants that decreased ADM significance after conditioning compared to those that did not (Mann-Whitney U *P*=5.4×10^−7^; **Fig. 4C**). These results remained significant even after adjusting effect sizes for standard error (Mann-Whitney U *P*=8.3×10^−3^; **Extended Data Fig. 8B**).

**Fig 4.**
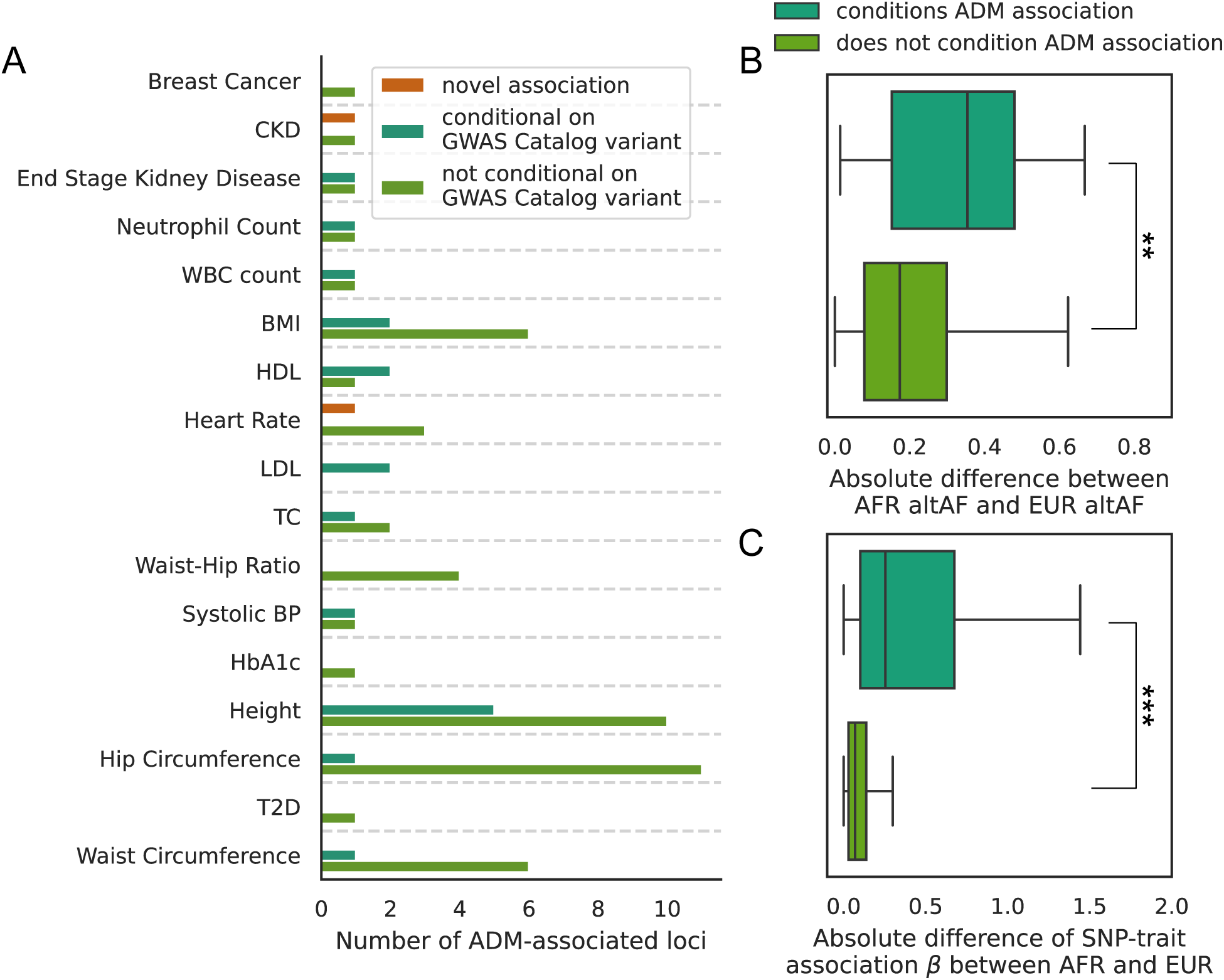
Characterization of ADM-associated loci. (A) Summary of conditional admixture mapping results, with the number of loci without any GWAS hits within 5Mb of the ADM loci (orange bar), number of loci with a GWAS hit that also reduces the ADM association (P>1×10^−4^) after conditional analyses (dark green bar), and the number of loci with a GWAS hit that does not reduce the ADM association (P<1×10^−4^) after conditional analyses (light green bar). (B) Boxplot comparing the difference in alternative allele frequencies (altAF) between 1KG EUR and AFR populations for variants which condition and do not condition ADM associations. (C) Boxplot comparing the difference in effect sizes from variant-trait associations in genetically-inferred AFR and EUR populations in All of Us for variants which condition and do not condition ADM associations. ** P<1×10^−3^; ***P<1×10^−5^

Unsurprisingly, the conditional variant with the largest difference in allele frequencies between AFR and EUR populations was rs2814778 within the Duffy locus, with an altAF of 0.97 in AFR populations and 0.01 in EUR populations. The altAF was also 0.98 in AFR local ancestry haplotypes and 0.005 in EUR local ancestry haplotypes as reported in gnomAD v4.1^39^. The conditional variant with the second largest difference in AF was rs1131877, which conditioned the association between loci 14q32.32 and BMI (**Fig. 5A**) and had an AFR altAF of 0.91 compared to an EUR altAF of 0.25 (**Fig. 5B**). The altAF was also 0.93 and 0.22 in gnomAD v4.1 AFR and EUR local ancestry haplotypes respectively. rs1131877 is a predicted missense variant within the gene *TRAF3*, which is also strongly associated with BMI in all individuals (Beta (95% CI)=0.13 (0.08,0.18); *P*=3.5×10^−7^), with similar effect sizes in AFR-like and EUR-like populations (**Fig. 5C**). *TRAF3* is involved in immune function and related disorders^40,41^, including metabolic dysfunction^42–44^. rs1131877 has also previously been nominated as explaining some variation between Hungarian and Anglo-Saxon populations in Brooke-Spiegler Syndrome symptoms and sarcoidosis risk in admixture analyses of self-identified African American men^45^.

**Fig 5.**
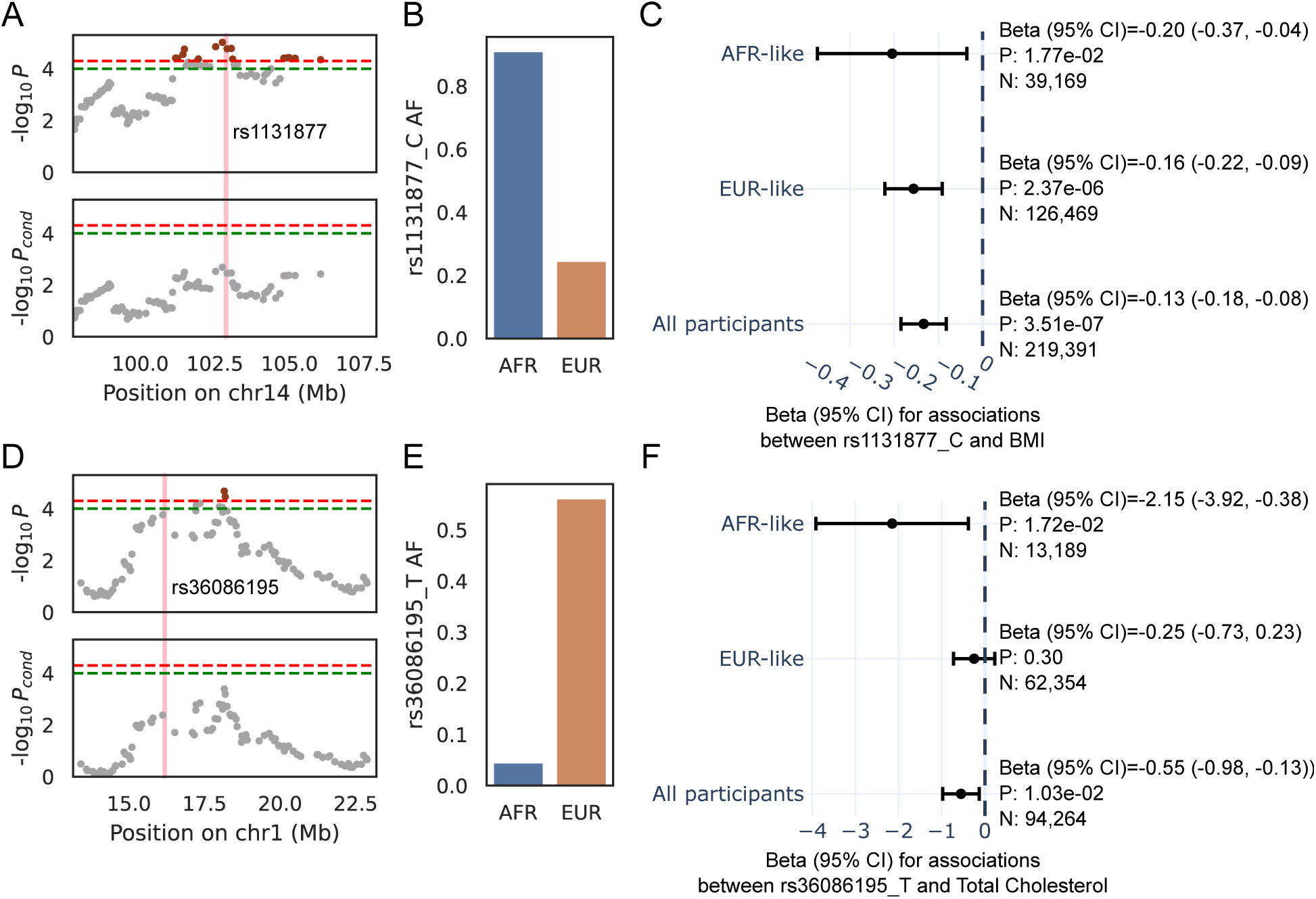
Differential allele frequencies and effect sizes underly ADM associations. (A) ADM association on chr14 with BMI, before and after conditioning on a previously published genome-wide significant variant from GWAS catalog (pink line). (B) Allele frequencies for the C-allele of rs1131877 in 1000G. (C) Effect sizes for associations of the C-allele of rs1131877 with BMI in AFR-like, EUR-like and all participants within AoU. (D) ADM association on chr1 with total cholesterol (TC), before and after conditioning on a previously published genome-wide significant variant from GWAS catalog (pink line). (E) Allele frequencies for the T-allele of rs36086195 in 1000G. (F) Effect sizes for associations of the T-allele of rs36086195 with TC in AFR-like, EUR-like and all participants within AoU.

The conditional variant with the largest effect size difference between AFR-like and EUR-like populations in AoU was rs36086195, which conditioned the association between 1p36.13 and total cholesterol (TC; **Fig. 5D**). Although rs36086195 is more common in EUR populations (AF=0.56) relative to AFR (AF=0.05; **Fig. 5E**), the effect size in AFR-like populations (Beta (95% CI)=-2.2 (-3.9,-0.4); *P*=0.02) was larger than that in EUR-like populations (Beta (95% CI)=-0.3 (-0.7,0.2); *P*=0.30; **Fig. 5F**). This could indicate differences in LD structure, statistical power, or differences in true causal effect sizes. Interestingly, the effect sizes are largely similar between AFR-like and EUR-like populations in Graham et al., which originally reported the association^46^, suggesting unique LD structures in AoU or non-genetic factors such as cohort differences. Graham et al. also performed fine-mapping on their multi-ancestry GWAS for TC, identifying three variants within the 95% credible set of this loci, including rs36086195. LD structure between these three variants differed greatly between AFR-like and EUR-like populations (rs36086195 vs rs924204: AFR-like R^2^=0.47, EUR-like R^2^=0.94; rs36086195 vs rs1497406: AFR-like R^2^=0.49, EUR-like R^2^=0.99; rs924204 vs rs1497406: AFR-like R^2^=0.82, EUR-like R^2^=0.94), suggesting LD differences between tagged and causal signals could explain the difference in observed effect size between populations. rs36086195 is a non-coding variant 1.9 Mb upstream of the lead ADM signal and is closest to the gene *EPHA2*, which encodes the ephrin type-A receptor 2, is involved in cell-cell signaling, and interacts with cholesterol^47–49^. rs36086195 is also associated with liver chromatin accessibility and alanine transaminase (ALT), highlighting the role of this locus in metabolic function^50^. In summary, these results demonstrate the utility of ADM to identify loci with different allele frequencies and heterogeneous effect sizes of variants that either are differentially tagging the causal signal or—less likely—are the causal signal itself.

### ADM uncovers novel genetic associations

The low number of ADM associations which decreased in significance after conditioning on a GWAS catalog variant suggests that our ADM results are tagging genetic signals which previous GWAS have not reported. To identify putative signals underlying these ADM associations, we performed single variant association analyses on all common variants within 10 Mb of an ADM association which could not be mapped to a published GWAS catalog signal, then re-ran ADM conditioning on all nominally significant variants (*P*<0.05; **Methods**). Thirty-two (60.3%) of ADM associations decreased in significance (*P*>1×10^−4^) after conditioning on a single variant (**Supplementary Table 11**). Interestingly, 26 (81.3%) of these variants did not reach genome-wide significance in the single variant association analyses, highlighting the utility of ADM to identify differentially frequent signals which traditional GWAS may miss. Twenty-seven of the variants were common (MAF>0.05) in either AFR or EUR 1000G populations, with 18 (66.7%) of these variants being more common in AFR compared to EUR populations (**Fig. 6A**; **Supplementary Table 11**). GWAS may have missed these variants due to the underrepresentation of individuals genetically similar to AFR populations. Indeed, the AFR-enriched variants were on average less significant in EUR-like GWAS compared to the 9 EUR-enriched variants (*P*=0.04; **Fig. 6B**). Similarly, these variants were also more significant in AFR-like and AFR-EUR admixed populations compared to the EUR-enriched variants, though these trends were not statistically significant likely due to limited sample sizes (**Fig. 6B**).

**Fig. 6.**
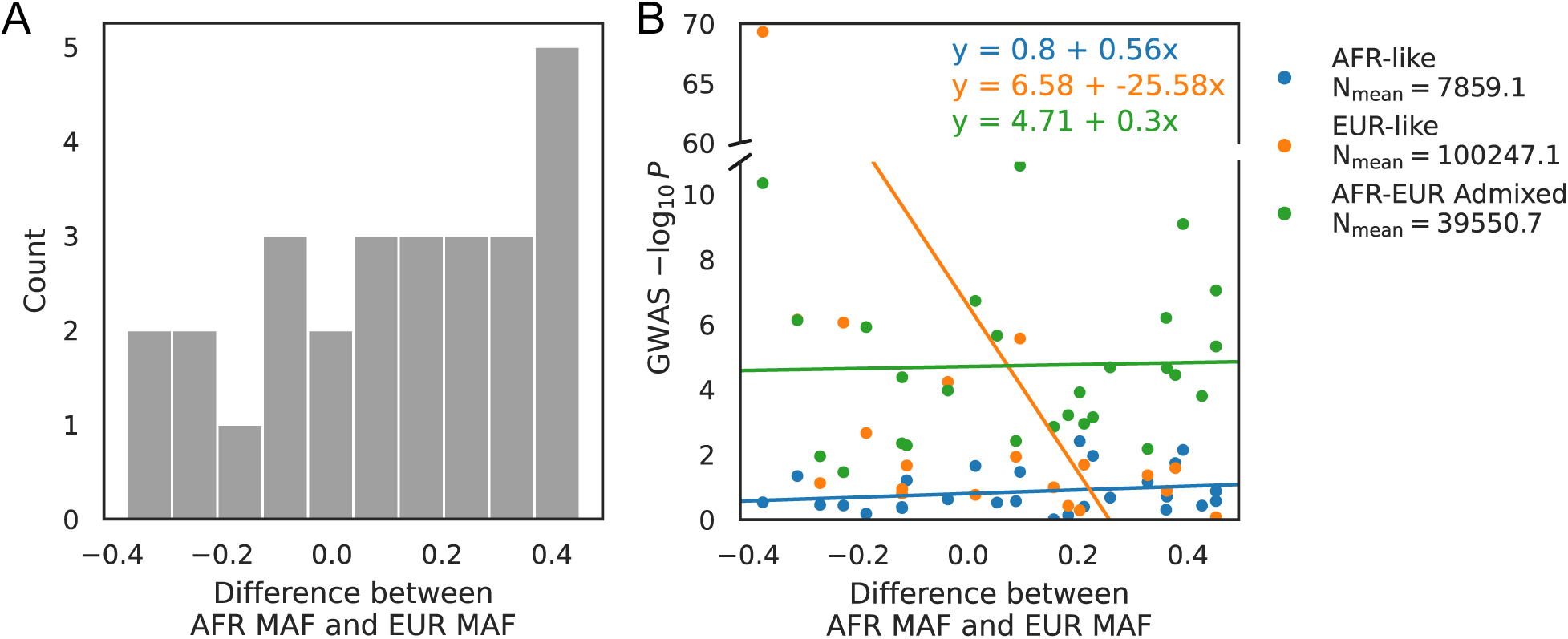
Novel ADM associations tag AFR-enriched variants. (A) Histogram of the difference in MAF between AFR and EUR 1000G populations for common variants which condition one of the ADM associations that cannot be attributed to a published association on GWAS catalog. (B) Comparison of the difference between AFR and EUR MAF values and the association significance of the variant from GWAS run within AFR-like, EUR-like, and AFR-EUR Admixed populations within AoU. Associations between MAF differences and GWAS significance are only significant for the EUR-like analysis (P=0.04).

As an example, one such novel ADM association was within region 9q21.33, where inferred local African ancestry was associated with increased risk of ESKD (OR (95% CI)=1.4 (1.2,1.7); *P*=3.6×10^−5^; **Fig. 7A, upper panel**). The closest published association in GWAS catalog was for the variant rs4744712, 15 Mb away from the lead loci, which did not change the significance of the ADM association after conditional analyses (*P*=3.7×10^−5^). Furthermore, this association remained significant even when including T2D status as a covariate (*P*=3.5×10^−5^), a common comorbidity of ESKD. Single-variant association analyses in AoU identified the variant rs56145118 as decreasing the ADM association significance the most after conditional analysis (*P_cond_*=5.6×10^−4^; **Fig. 7A, lower panel**). This variant is located within the intron of *SLC28A3* and the first exon of the non-coding RNA gene ENSG00000285987 (**Fig. 7B**). The insertion of AAACAACAACAAC is more common in AFR populations (AF: 0.23) relative to EUR populations (AF: 0.001) (**Fig. 7C**), with increased ESKD risk for the insertion (OR (95% CI)=1.3 (1.1,1.4); *P*=7.1×10^−4^; **Fig. 7D, upper panel**), matching the increased risk with AFR local ancestry haplotypes observed in ADM. To further characterize this variant, we calculated its association with related quantitative traits, and similarly found significant associations with standardized eGFR (Beta (95% CI)=-0.02 (-0.04,-0.01); *P*=5.6×10^−4^) and creatinine (Beta (95% CI)=0.04 (0.02,0.06); *P*=9.4×10^−4^), as well as a borderline significant association with BMI (Beta (95% CI)=-0.01 (-0.02,0); *P*=0.06; **Fig. 7D, lower**). While little is known on the function of ENSG00000285987, *SLC28A3* is a sodium-dependent nucleoside transporter with high protein levels in the kidney^51^. Previous rare variant association analyses have also reported moderate associations between *SLC28A3* and ESKD^52^, and *SLC28A3* is overexpressed in kidney plasma cells of patients with chronic kidney disease (CKD)^53^. To explore if rs56145118 may be tagging other exonic variants, we calculated R^2^ between rs56145118 and all other exonic variants proximal to rs56145118 (**Methods**). We observed low correlation with the other exonic variants, with the most correlated variant being in the 3’UTR of *SLC28A3* with an R^2^ of 0.1 in both AFR and EUR local ancestry haplotype homozygous carriers (**Supplementary Table 12**).

**Fig. 7.**
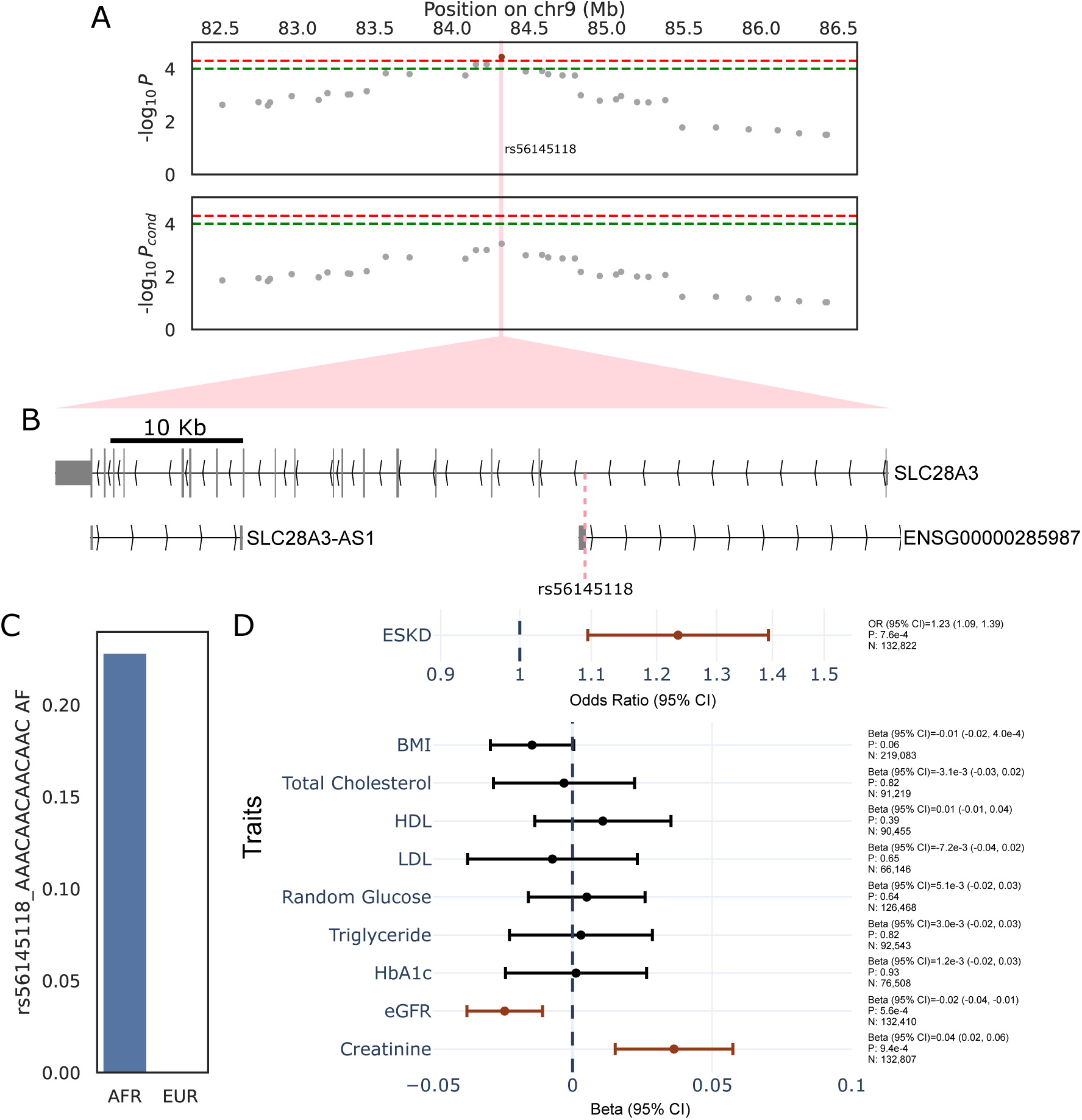
SLC28A3 variation is associated with End-Stage Kidney Disease. (A) Upper Panel: Novel End-Stage Kidney Disease (ESKD) associated loci identified with ADM, with increased risk of ESKD associated with inferred local African ancestry. The red dotted line represents the genome-wide significance threshold (P<5×10^−5^) and the green dotted line represents the suggestive significance threshold (P<1×10^−4^). Lower Panel: The ADM association decreases in significance after conditioning on the variant rs56145118. The vertical pink line represents the position of rs56145118. (B) rs56145118 is located within the first intron of SLC28A3 and the exon of the non-coding RNA gene ENSG00000285987. (C) Allele frequencies for the AAACAACAACAAC insertion of rs56145118 in 1000G. (D) Forest plots of odds ratios for ESKD and standardized effect sizes of related quantitative traits for rs56145118, relative to the AAACAACAACAAC insertion, across all individuals in All of Us. Red points indicate nominally significant (P<0.05) associations.

## Discussion

Here, we aimed to identify genetic loci with differential enrichment and effects between ancestry groups to identify novel disease associations. To this end, we performed the largest admixture mapping efforts to-date in AFR-EUR Admixed individuals, identifying 59 loci where differences between AFR and EUR local ancestries are associated with at least one of 22 different quantitative and binary traits, for a total of 71 trait-to-loci mappings. Importantly, comparison of ADM-identified signals with known GWAS associations revealed that 75% of ADM associations were independent of variants from a published GWAS. Sixty percent of these independent associations could be linked to a single variant in AoU, a majority of which were enriched in AFR populations. Similar to IBD-mapping associations which can identify rare variant associations missed in traditional GWAS^54^, these ADM signals may represent novel disease-associated loci which previous GWAS have not reported due to reduced power from the limited representation of participants genetically similar to African populations. Consequently, they may include novel biological insights into diseases to identify treatment options and improve disease prediction models. Furthermore, 81% of these associations did not reach the traditional GWAS genome-wide significance threshold, highlighting the power of admixture mapping for identifying associations which GWAS may miss.

Beyond these association analyses, investigations of local ancestry frequencies within admixed individuals can shed light on regions under selection since admixture. Using a sample size almost 20K larger than the previous largest such investigation of selection in AFR-EUR Admixed individuals^33^, we found evidence of selection at the HLA locus. Previous studies have similarly described enrichment of African ancestry at the HLA locus, with improvements in immune function from these African-ancestry HLA alleles potentially driving selection^55–57^. We also found similar patterns of enrichment in a smaller replication sample, even when masking the HLA locus, suggesting errors in local ancestry inference and sequencing at the locus may not be solely driving this trend. Together, these results highlight possible rapid selection of immune-relevant genes due to admixture between African and European populations. However further work is required to determine if the observed enrichment of AFR local ancestry haplotypes at the HLA locus is truly due to selection rather than other factors such as other sources of errors in local ancestry inference methods.

Previous work has demonstrated the utility of admixture mapping for identifying loci with differing causal variant allele frequencies between populations^27^, which we also confirmed when comparing altAF values across variants which reduce ADM significance after conditional analysis. Furthermore, we found evidence that effect sizes also vary for these variants when estimated in predicted African and European populations. As causal effects are mostly shared between populations^25^, these differences may represent large standard errors due to differences in power from allele frequency variations, differential LD architecture with true causal signals, or exceptions to the largely shared effect-sizes. Interactions between genetics and non-genetic factors could also explain some of these differences, but further work is required to determine if such interactions exist at these loci.

Disease prevalence and trait distribution are known to vary across populations. For example African Americans have a higher prevalence of ESKD compared to European Americans, motivating ADM studies in African American participants^58^. In previous ADM studies, increased ESKD risk was associated with African ancestry near the *APOL1* locus, ultimately leading to the development of drugs targeting *APOL1* variations^59^. Thus, ADM results not only can help explain prevalence variations between populations, but also lead to novel therapeutic options for patients. In this study, we identified a novel association between inferred local African ancestries at the *SLC28A3* locus and increased risk of ESKD, which an intronic variant within *SLC28A3* may partially explain. Such results identify an additional source of ESKD risk and may lead to novel therapeutic approaches to preventing ESKD progression. As this variant is both in the intron of *SLC28A3* and the first exon of a non-coding RNA gene, further work is required to determine the biological mechanisms, and additional underlying variants, of this association.

We conclude with several limitations. First, heterogeneity within our genetically-inferred AFR-EUR Admixed samples could potentially result in non-genetic effects being differentially present in some individuals, leading to confounders driving some of our observed associations. Second, we performed LD clumping on ADM results to identify lead loci. However, the high correlation of local ancestry segments could result in loci lacking a causal variant to be identified as a lead loci. Consequently, we generated broad boundaries around lead loci for GWAS signal identification. We anticipate that future methods improving fine mapping efforts on ADM results can help parse these results. We also recognize the minimal overlap of significant ADM associations in our replication efforts, especially for UKBB. Low statistical power may underly the low replication, evident from the high effect size correlation between the replication ADM and AoU ADM results. However, this low overlap is also expected due to the differences in genetic architectures and population histories of AFR-EUR Admixed populations in the US and UK^60,61^, as well as other non-genetic differences between AoU, an EHR-based biobank, and UKBB, a population-based biobank. Finally, we only performed conditional analyses with one variant at a time. However, multiple variants could jointly underly ADM associations and such interplays are not considered in our analyses. As we tested thousands of variants in our single-variant conditional analysis models, adding multiple variants per model was computationally infeasible.

In summary, we performed the largest local ancestry inference effort in the *All of Us Research Program*. Leveraging this data, we identified loci under putative selection since admixture within AFR-EUR admixed individuals and found novel associations from ADM which have not been described in GWAS. As inclusion of recently admixed individuals in genetic studies increases, these results suggest ADM efforts will continue to identify novel genetic associations and insights into trait biology.

## Extended data figures

**Extended Data Fig. 1.**
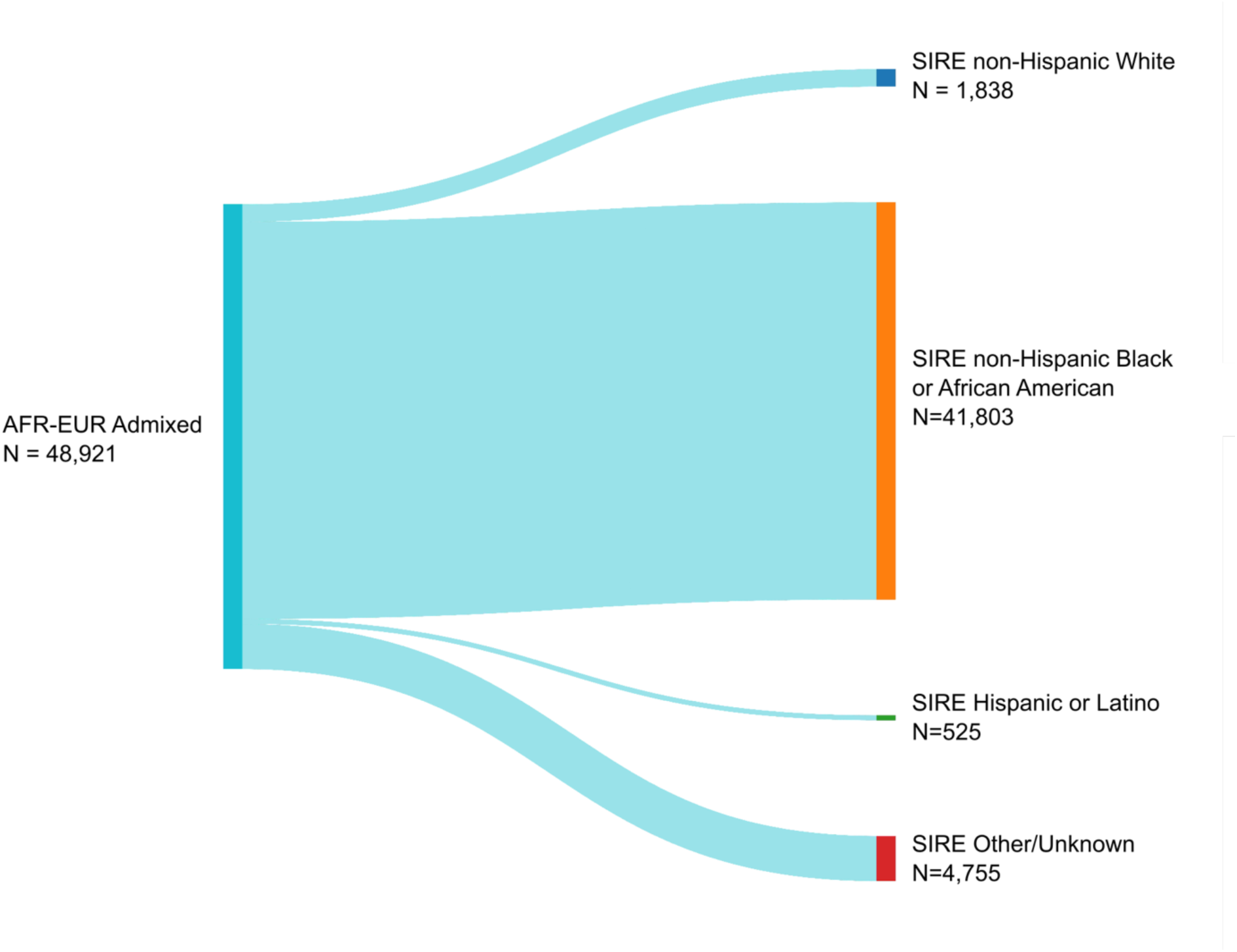
Overlap of genetically inferred AFR-EUR Admixed individuals with SIRE categories. A Sankey plot of AFR-EUR Admixed individuals and their self-reported race and ethnicity.

**Extended Data Fig. 2.**
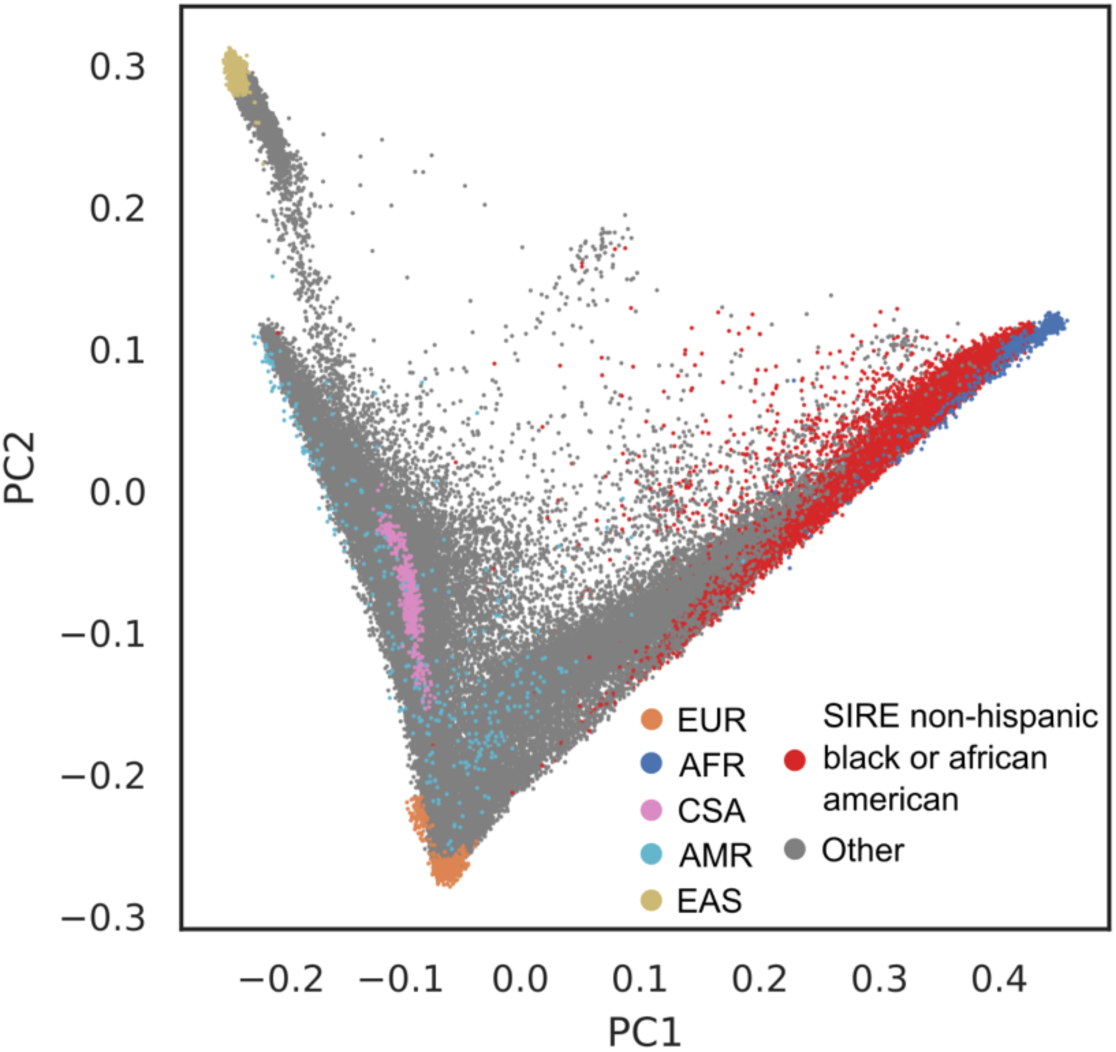
Overlap of SIRE non-Hispanic Black or African American participants in AoU and reference populations from 1000G. AoU participants plotted in PC space, along with reference populations from 1000G. AoU participants who are not self-report as being non-Hispanic Black or African American colored grey.

**Extended Data Fig. 3.**
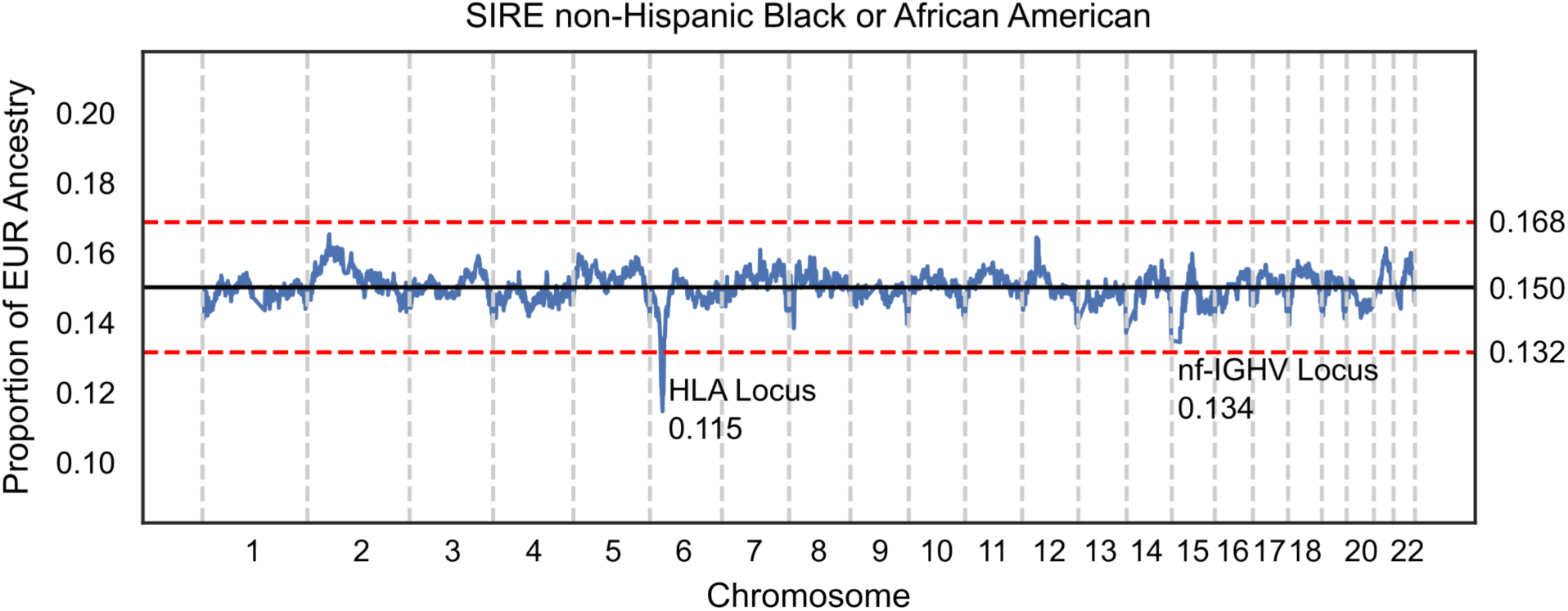
Putative evidence of selection since admixture in SIRE non-Hispanic Black or African Americans. Proportion of local EUR ancestry across the genome in unrelated, self-reported non-Hispanic Black or African American participants in All of Us. The solid black line represents the genome-wide average proportion of local EUR ancestry, and the red dashed lines represent the P<0.05 cutoff of 4.2 SD away from the mean.

**Extended Data Fig. 4.**
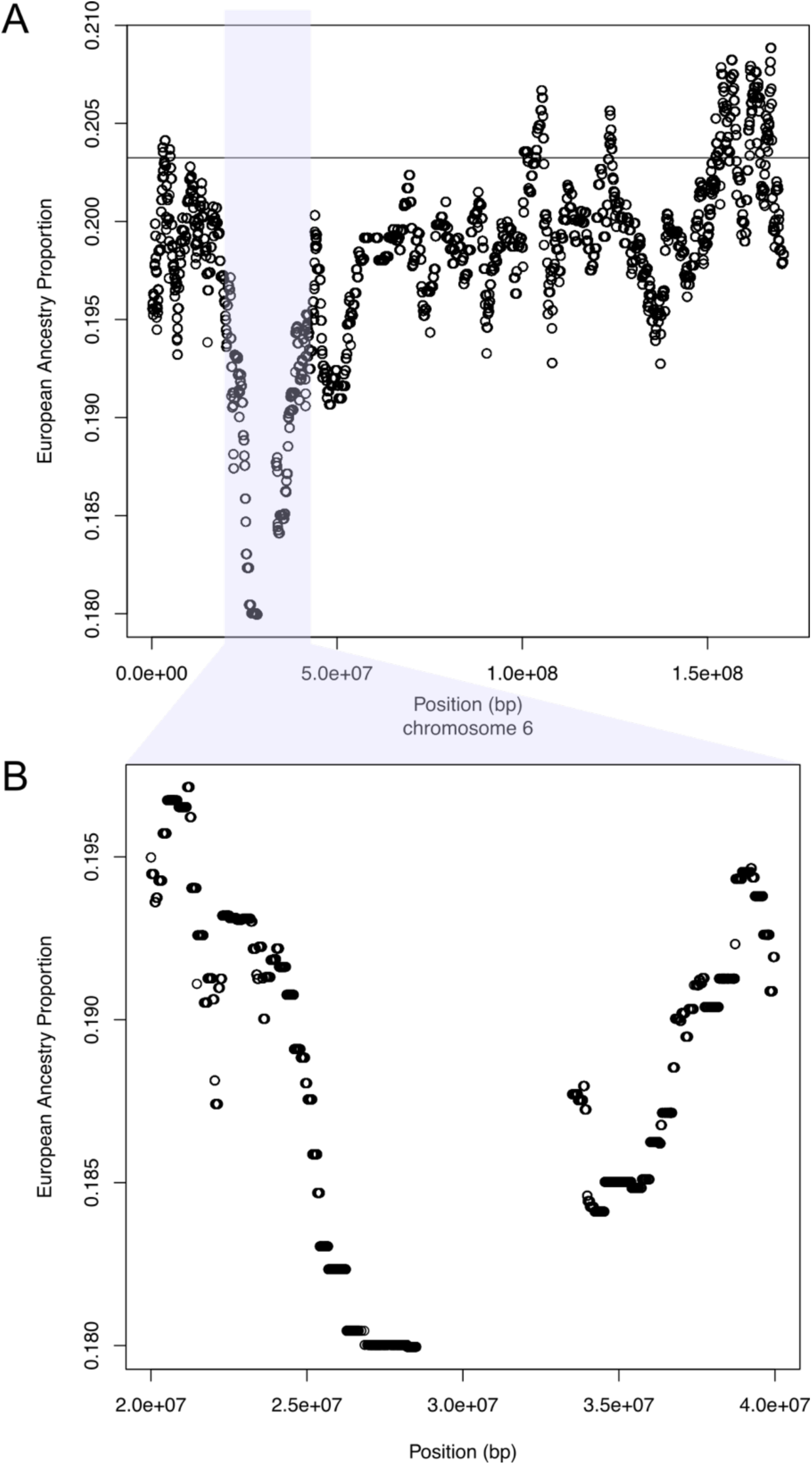
Putative evidence of selection since admixture in PMBB. (A) Proportion of local EUR ancestry across chromosome 6 in AFR-EUR Admixed participants within PMBB. The solid black line represents the genome-wide average proportion of local EUR ancestry. (B) Proportion of local EUR ancestry around the HLA locus. Local ancestry inference in both (A) and (B) was performed after masking the HLA locus.

**Extended Data Fig. 5.**
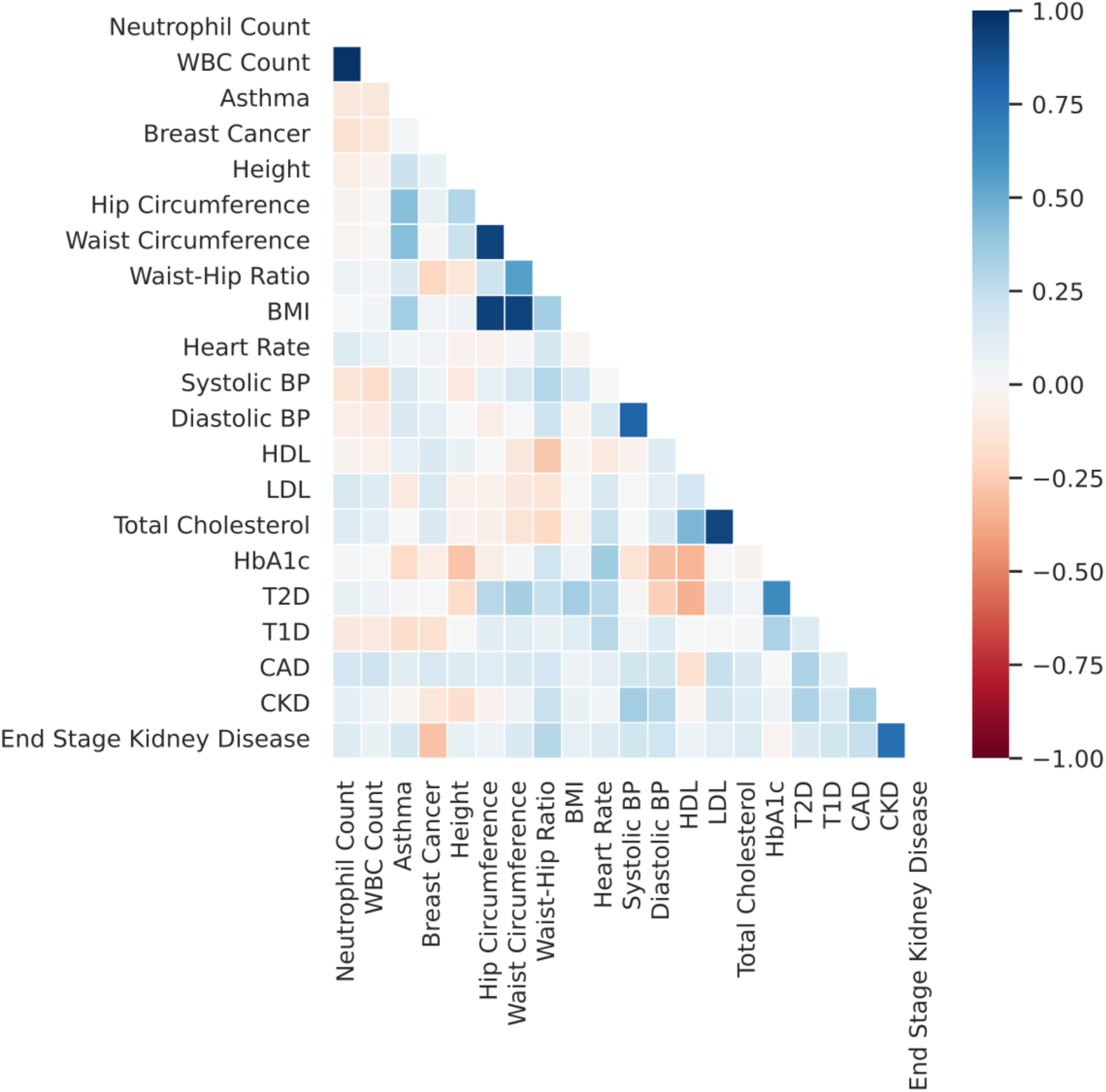
Correlation of effect sizes across ADM-associated loci. Plotted is a heatmap of Pearson R correlation values between effect sizes per trait pair across all independent loci with at least one genome-wide significant ADM association.

**Extended Data Fig. 6.**
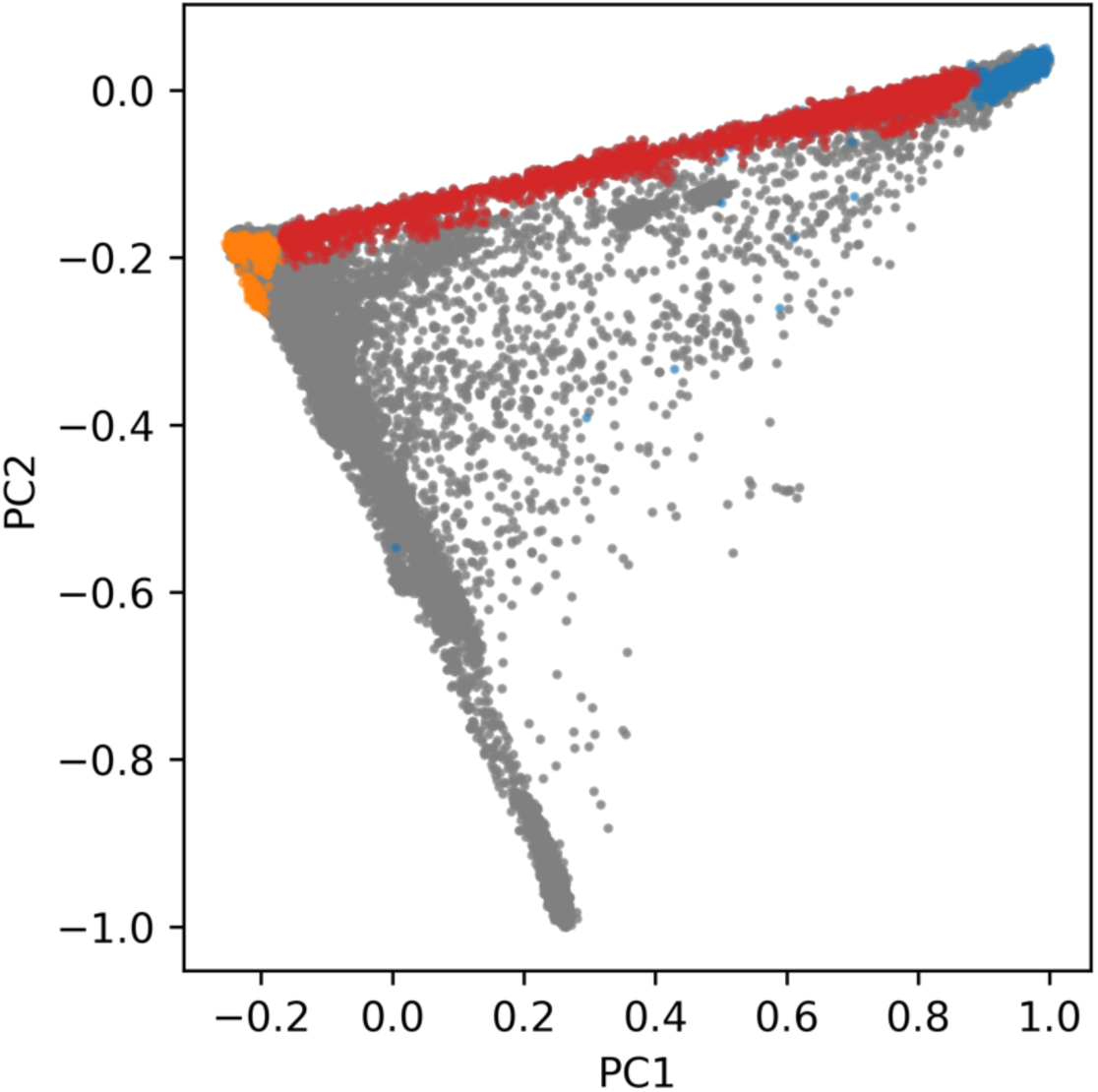
UK Biobank participants plotted in PC space. Red dots highlight genetically inferred UK Biobank two-way admixed AFR/EUR individuals, gray dots denote the entire UK Biobank population, orange dots denote 1000G EUR individuals, and blue dots denote 1000G AFR individuals.

**Extended Data Fig. 7.**
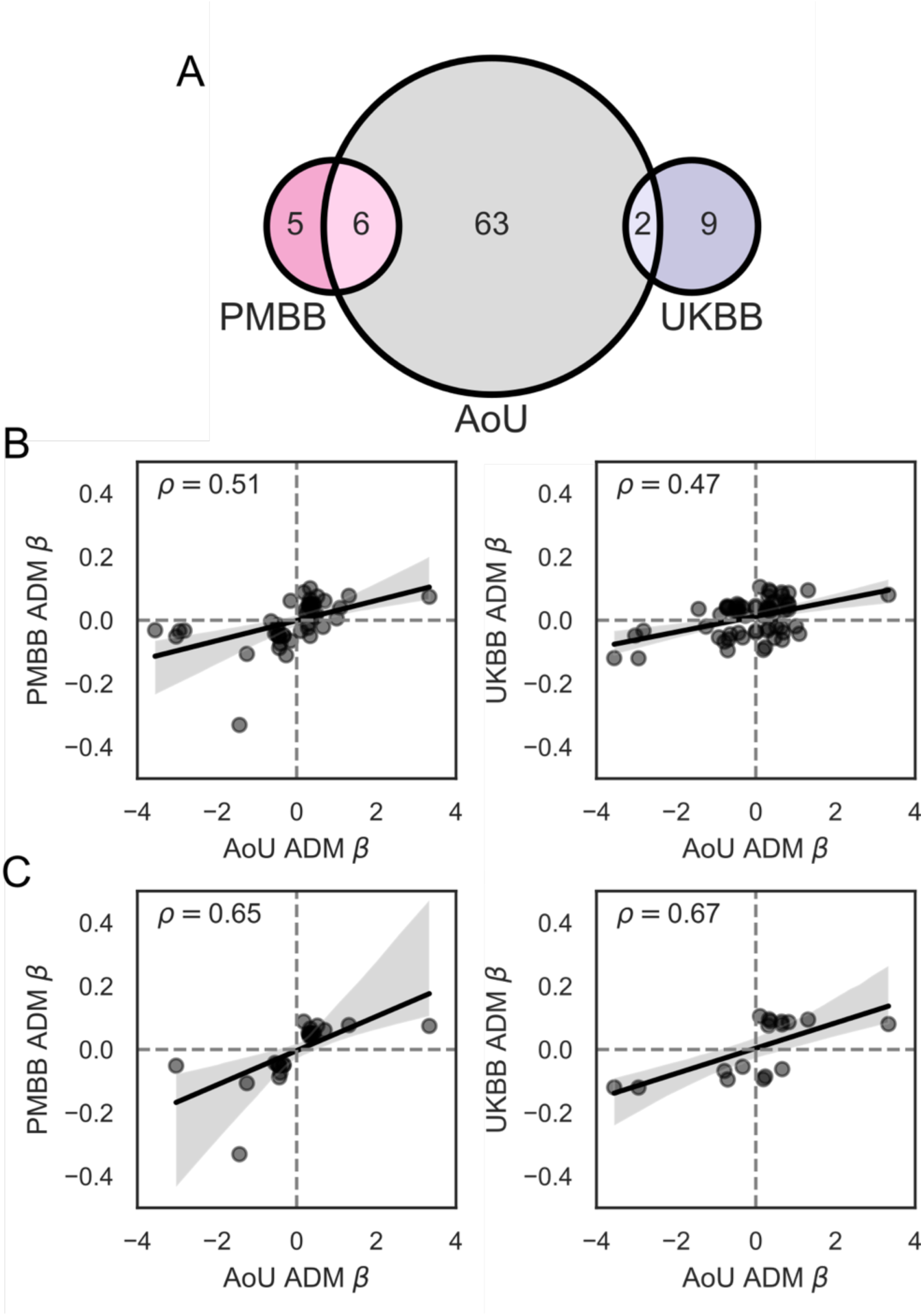
Comparison of admixture mapping results between All of Us, UK Biobank, and Penn Medicine Biobank. (A) Venn diagram of the overlap of genome-wide significant admixture mapping (ADM) associations across All of Us (AoU), UK Biobank (UKBB), and Penn Medicine Biobank (PMBB). (B) Scatterplots of ADM effect sizes between AoU and UKBB and between AoU and PMBB. (C) Scatterplots of ADM effect sizes between AoU and UKBB for ADM associations with P<0.05 in UKBB and between AoU and PMBB for ADM associations with P<0.05 in PMBB.

**Extended Data Fig. 8.**
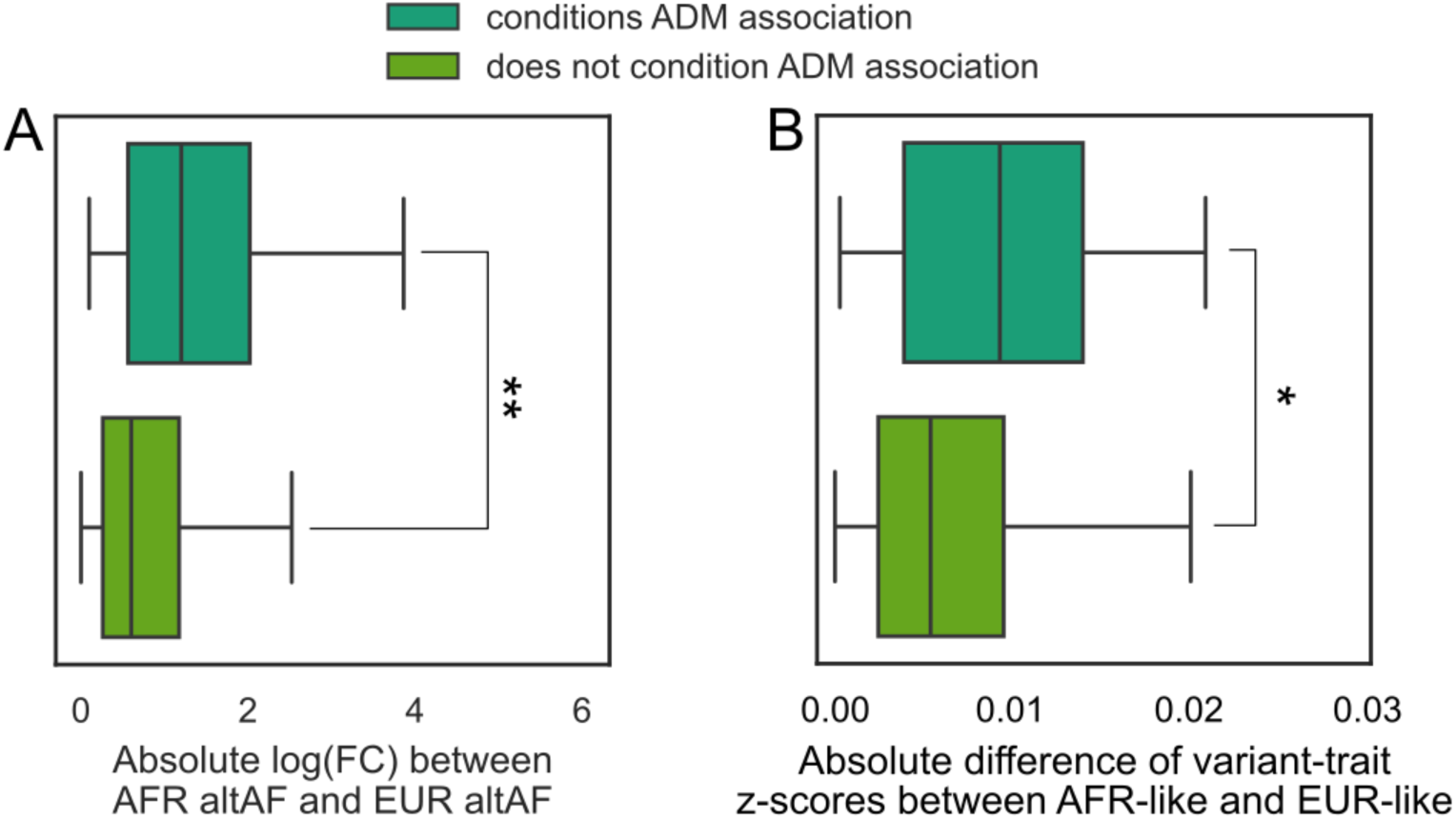
Alternative comparisons of variants which condition ADM associations. (A) Comparison of absolute log fold-change (FC) for the same alternative allele frequencies (altAF) between AFR and EUR populations from 1000G for variants which condition and do not condition ADM associations. (B) Comparison of absolute difference in z-scores for variant-trait associations between AFR-like and EUR-like populations for variants which condition and do not condition ADM associations. **P<1×10^−3^. *P<0.01

## Methods

### Overview of cohorts

#### All of Us Research Program

The All of Us Research Program (AoU) is a large, US-based biobank developed to improve and enable the inclusion of diverse, underrepresented populations in large-scale genetic and epidemiological research studies^13,32^. We accessed short read whole genome sequencing (srWGS) data, survey data, electronic health record data from participants included in the v7 release of AoU. Age within AoU was calculated as participants’ age as of August 1, 2022. Sex was determined using self-reported sex-at-birth data provided by AoU. We identified individuals as being non-Hispanic Black or African American if their self-reported race was “Black or African American” and their self-reported ethnicity was “not Hispanic or Latino”.

AoU provides a list of flagged individuals with poor srWGS data quality based on the number of variants, insertion/deletion ratios, number of insertions, number of deletions, heterozygosity of variants or indels, transition/transversion ratios, and number of variants not in gnomAD v3.1. Additionally, variants from srWGS were flagged as being low quality based on a GQ threshold of <20, a DP threshold of <10, an allele balance filter of <0.2, a low QUAL score (60 for SNPs and 69 for indels), or an ExcessHet score <54.69. These individuals and variants were removed from all subsequent analyses. AoU additionally provides a minimized list of related individuals, with relatedness determined using PC-Relate to account for population structure when calculating relatedness, which was used to remove related participants^62^.

To infer local and global ancestry in AoU, we first extracted genetic data from the 1000 Genomes Project (1000G) for individuals from each superpopulation. We then calculated principal components within these individuals, then projected the AoU participants into the same PC space. These PCs were additionally used in all statistical models in AoU. Finally, we estimated inferred local and global ancestry with RFMix v2^63^ with default parameters, using the 1000G data above as reference and HapMap3 variants. AFR-EUR admixed individuals were identified if their combined global ancestry proportions of AFR and EUR was >0.95 but the individual proportions of AFR and EUR was <0.95. This definition was used to identify individuals with any level of admixture between African and European populations.

#### UK Biobank

The UK Biobank (UKBB) is a large-scale biomedical database containing genetic and electronic health record information from half a million residents from the United Kingdom. Genotype data was prepared as described in Hou et al. 2023^25^. Briefly, pre-imputed variants were retained with imputation R² > 0.8, minor allele frequency (MAF) > 0.5%, and ancestry-specific MAF > 0.5% in both the African and European populations. Ancestry inference was performed using SCOPE v6^64^, which estimated global ancestral proportion based on 1000G allele frequencies, where 4,326 individuals with more than 5% of both AFR and EUR ancestries. Furthermore, local ancestry was inferred using RFMix v2 with default setting using 99 Northern and Western European residents in Utah (CEU) in 1000G and 108 Yoruba individuals from Ibadan, Nigeria (YRI) in 1000G. Only HapMap3 SNPs were used for local ancestry inference, with other variants interpolated.

#### Penn Medicine Biobank

The Penn Medicine Biobank (PMBB) collects genetic data and electronic health record data from consenting individuals within the University of Pennsylvania Health System. Zaidi et al. previously reported local ancestry inference within PMBB participants genetically similar to AFR^37^. Briefly, genotype data was phased using Beagle then local ancestry inference was performed using RFMix v1 with CEU and YRI populations as reference. The HLA locus was masked before local ancestry inference.

### Phenotypes

We analyzed a total of 7 binary and 15 quantitative traits in AoU, 5 binary and 16 quantitative traits in the UKBB, and 7 binary and 12 quantitative traits in PMBB. For the AoU dataset, we selected the most recent measurements categorized under “condition” and “lab and physical measurement,” identified using “concept ID” and ICD-10 codes for the binary traits. A full description of the phenotypes selected has been described previously^65^. Outliers in the quantitative traits, defined as values exceeding 5 standard deviations from the mean, were excluded from the analysis. In UKBB, traits with corresponding definitions and ICD codes were extracted following the procedures outlined in the script available at https://github.com/privefl/UKBB-PGS/blob/main/code/prepare-pheno-fields.R. In PMBB, quantitative and binary traits were derived from patients’ electronic health records.

### Admixture mapping

Association of local ancestry haplotypes with phenotypes was performed using admix-kit with the ADM method^66^. Associations in all cohorts were adjusted for age, sex, and the first 10 principal components. Additionally, models generated within AoU additionally adjusted for the site where the srWGS was performed. Single ancestry haplotype analyses were performed for AFR-EUR Admixed individuals within AoU and UKBB, where the effect of having an AFR haplotype relative to not having an AFR haplotype within a specific local ancestry locus on phenotypes was assessed. For BMI, we compared the associations we detected with ADM to those from AoU GWAS. A full description on the GWAS can be found here (https://support.researchallofus.org/hc/en-us/articles/27049847988884-Overview-of-the-All-by-All-tables-available-on-the-All-of-Us-Researcher-Workbench). Briefly, GWAS was run using SAIGE^67^ on quantitative traits and phecodes within AoU, stratified by genetically inferred ancestry groups. These genetically inferred ancestry groups were determined using a random forest classifier trained on data from gnomAD, 1000G, and the Human Genome Diversity Project^32^.

To determine the effective number of tests to adjust for, we explored three different approaches. First, we used an MCMC approach previously described^68^, and estimated an average of 297 tests. Second, we used the number of local ancestry segments in high LD with each other, using PLINK with a window or 5 Mb and R2 cutoff of 0.8, and estimated a total of 757 tests. Finally, we explored a cutoff of 1000 tests from literature searches of previous admixture mapping efforts^69,70^. We opted to use the latter approach to minimize the number of false positives, as well as allow for a constant significance threshold across the replication datasets.

After generating ADM summary statistics, loci were clumped to find independent significant hits, using the PLINK clumping function with significance threshold of 5 × 10^−5^, or an adjusted P<0.05, LD threshold of 0.1, and physical distance threshold of 5Mb at each chromosome for every trait. LD reference panels were constructed using inferred local AFR haplotypes from unrelated AFR-EUR Admixed individuals per local ancestry segment.

As local ancestry inference errors are concentrated in low-complexity regions^36^, we tested whether our ADM hits overlap these regions. Since there are many small low-complexity regions from the UCSC genome browser, and the local ancestry segments are large, 90.1% of the genome-wide significant ADM associations intersected with at least one low-complexity region. The amount of overlap was minimal, ranging from 0.05% to 1.80% (**Supplementary Table 5**). Furthermore, 5 of the ADM associations contained one HapMap3 variant used for local ancestry inference which was in a low complexity region. However, the total number of input HapMap3 variants used for each of these 5 regions ranged from 26 to 116, meaning only a small percentage of these input variants could have created errors stemming from the low-complexity regions (**Supplementary Table 5**). Since these results suggest that errors in local ancestry inference due to low-complexity regions may not greatly impact our results, we did not perform any filtering by low-complexity regions.

To identify if any genome-wide significant loci are the same across traits, we checked if any of the genome-wide significant loci from the 71 associations are in correlated with each other (R2>0.8). In total, this left us with 52 loci which are genome-wide significant for an ADM association and independent from other loci. For the 12 loci associated with multiple traits, we tested for evidence of colocalization using COLOC 5.2.3^71^ and a posterior probability of colocalization (PP.H4) >0.8. Colocalization boundaries were calculated with a 10 Mb window around the center of the local ancestry segment. This large window was selected to ensure enough local ancestry haplotypes were included in the colocalization analysis. We calculated pleiotropy of effect sizes across all ADM associations using z-scores calculated for each of the 22 traits at the 52 independent loci. We used the equation Z = B / (sqrt(N) * SE) to calculate z-scores per trait. We then calculated Pearson correlations between Z-scores per trait across the 52 loci.

We identified shared loci between the AoU and UKBB and PMBB ADM analyses by looking for intersections of local ancestry segments with genome-wide significant ADM associations for the same trait in both datasets.

### Evidence of selection

Inferred local EUR frequencies for unrelated AFR-EUR Admixed individuals and SIRE non-Hispanic Black or African American individuals were calculated using PLINK. We removed the first and last 2 Mb of each chromosome to avoid errors with local ancestry inference in low quality genomic regions.

We tested for associations between inferred local EUR ancestry and any phecode or relevant immune lab using the same covariates described above for ADM. P-values for binary and quantitative traits were independently FDR adjusted using the Benjamini-Hochberg procedure with a cutoff of 0.05.

### Conditional ADM analyses

To perform the admixture mapping analysis conditional on genotype variants, we first collected GWAS associations of from the GWAS catalog for the same traits with an ADM association and converted them to hg38 using liftOver. We then removed variants which were farther than 20 Mb from any significant ADM association. We used a large window around the ADM associations to account for the high correlation of local ancestry haplotypes. To avoid testing duplicate signals, we further filtered variants using a clumping strategy, with a MAF cutoff of 0.01, P-value threshold of 5×10^−8^, LD threshold of 0.01, and distance threshold of 5×10^6^. For each trait, we re-ran admixture mapping, additionally including an LD-independent genome-wide significant variants as a covariate. We ran these conditional models for each genome-wide significant variant such that no model included more than one variant. We determined whether a variant is involved in the ADM association if the ADM P-value dropped below 1×10^−4^, or a P-value greater than 0.1 after adjusting for 1000 different tests. To avoid cases where missing variants drops the number of samples such that decreases in the P-value could instead be due to power issues, we only considered conditional analyses where the sample size was at least 95% of that of the original analysis.

### Single variant association analyses

We calculated allele frequencies of variants of interest using data from 1000G continental populations labelled as EUR or AFR.

We estimated effect sizes for single variants with their traits of interest in AoU using either linear or logistic regression models depending on the trait type. Variants within 10 Mb of an ADM peak were extracted from the AoU ACAF files, which are pre-filtered to include only variants with a MAF >1% in at least one ancestry superpopulation. All models were adjusted for age, sex, the first 10 PCs, and sequencing site. Models were run in AoU participants genetically-similar to AFR 1000G populations (AFR-like), AoU participants genetically-similar to EUR 1000G populations (EUR-like), or all AoU participants. We defined AFR-like participants as those with a global AFR proportion >0.9, and EUR-like participants as those with a global EUR proportion >0.9.

When evaluating the significance of difference in allele frequency and effect size variations between populations for variants which condition and do not condition an ADM association, we utilized the non-parametric Mann-Whitney U Test, since we were comparing the absolute difference values, which are not normally distributed.

### Single variant fine-mapping

To determine if any specific variants may be underlying the ADM association which we could not assign to a known GWAS loci, we ran clumping on the single variant associations generated using all AoU participants with a P-value threshold of 0.05. LD threshold of 0.01, and distance threshold of 5×10^6^. We then re-ran the conditional analyses pipeline described above on all lead variants from this clumping step.

To identify additional variants which may underly the ADM association with ESKD at chromosome 9, we extracted exonic variants from all genes flanking rs56145118, including *SLC28A3*, *NTRK2*, *SLC28A3-AS1*, and ENSG00000285987, and calculated R^2^ in AFR-like and EUR-like populations using PLINK.

## Author Contributions

R.M. performed ADM, selection scans, and single variant analyses within AoU, analyzed the results, and led the study design and manuscript writing. Z.S. performed admixture mapping within PMBB and UKB, integrated ADM associations between datasets, ran conditional analyses using GWAS catalog variants, and contributed to the writing. K.H. ran local ancestry inference and contributed to study design. Y.W. prepared AoU phenotype data. G.M. tested for replication of selection signal in PMBB. A.J.A contributed to ADM replication analyses in PMBB. S.C., E.K., E.A., and A.R.M. provided feedback throughout the project and revised the paper. B.P. conceived the study, supervised the analyses, and contributed to the manuscript writing.

## Acknowledgements

We gratefully acknowledge *All of Us* participants for their contributions, without whom this research would not have been possible. We also thank the National Institutes of Health’s [All of Us Research Program](https://allofus.nih.gov/) for making available the participant data examined in this study.

We also acknowledge the Penn Medicine BioBank (PMBB) for providing data and thank the patient-participants of Penn Medicine who consented to participate in this research program. We would also like to thank the Penn Medicine BioBank team and Regeneron Genetics Center for providing genetic variant data for analysis. The PMBB is approved under IRB protocol #813913 and supported by Perelman School of Medicine at University of Pennsylvania, a gift from the Smilow family, and the National Center for Advancing Translational Sciences of the National Institutes of Health under CTSA award number UL1TR001878.

## Data Availability

Individual-level phenotype and genetic data from All of Us are available on the All of Us Research Workbench for all approved, controlled-tier researchers, and phenotype and genetic data from UK Biobank are also publicly available for approved researchers. Inferred local ancestry data can be made available to other approved All of Us and UK Biobank researchers upon request. Individual-level data from Penn Medicine BioBank cannot be shared to researchers without an approved Penn Medicine BioBank IRB, but summary statistics can be made available upon reasonable request.

