## Supplementary figures and images for "Large-scale admixture mapping in the *All of Us Research Program* improves the characterization of cross-population phenotypic differences"

### Extended Data Fig. 1

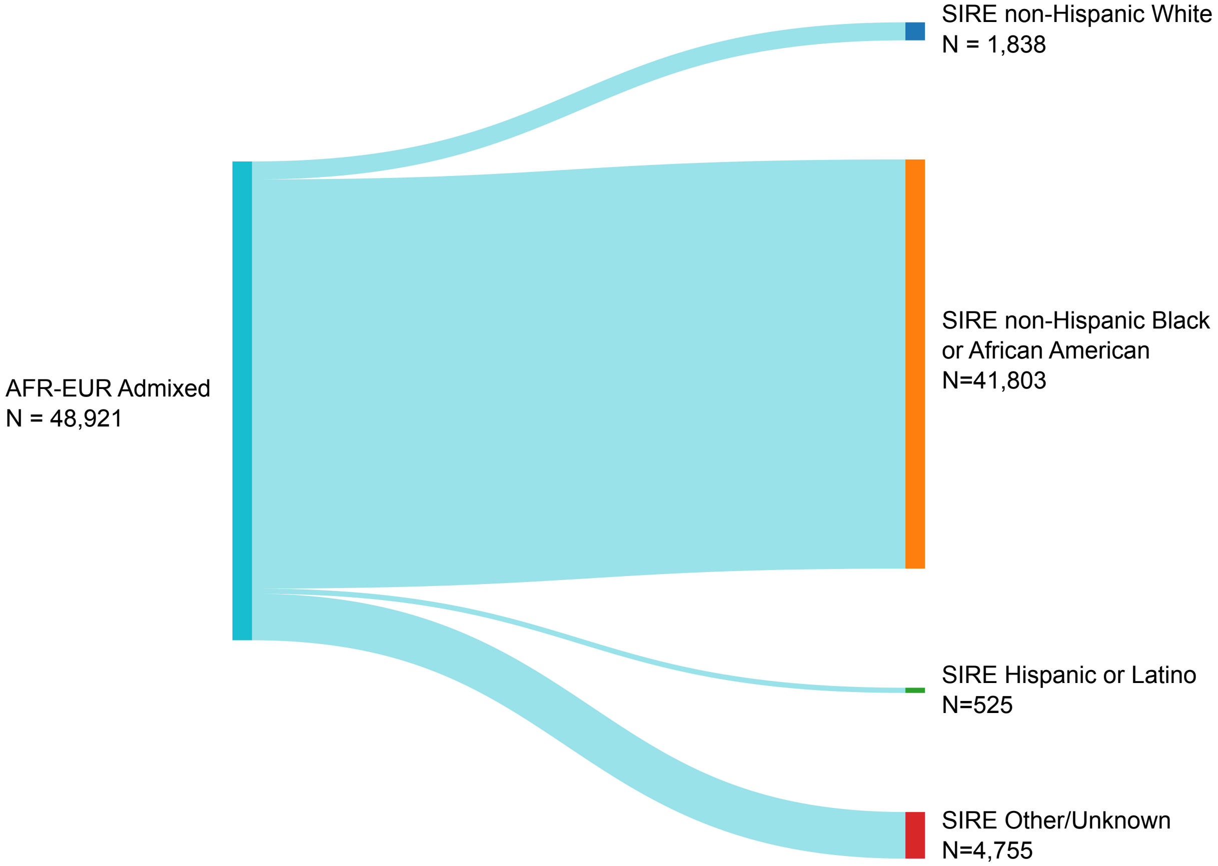

### Extended Data Fig. 2

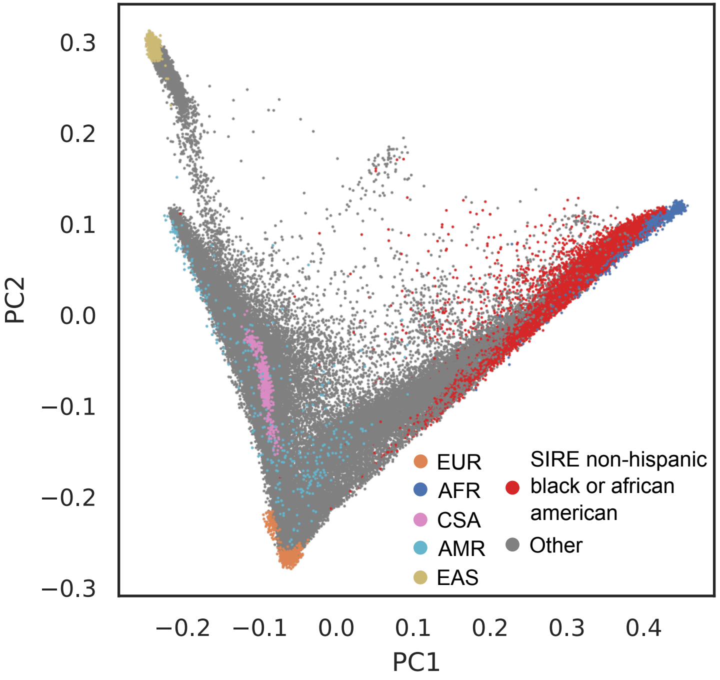

### Extended Data Fig. 3

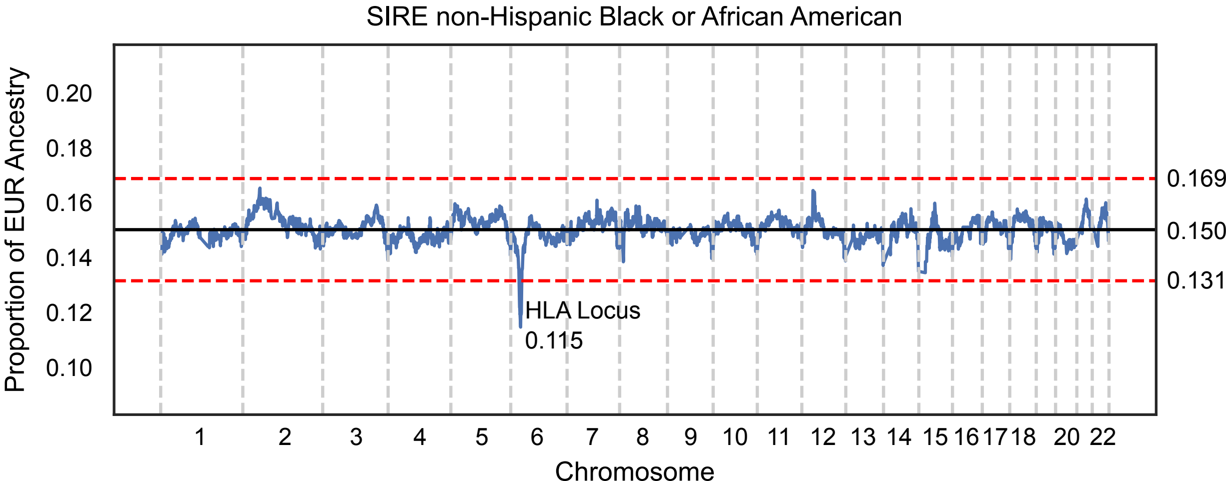

### Extended Data Fig. 4

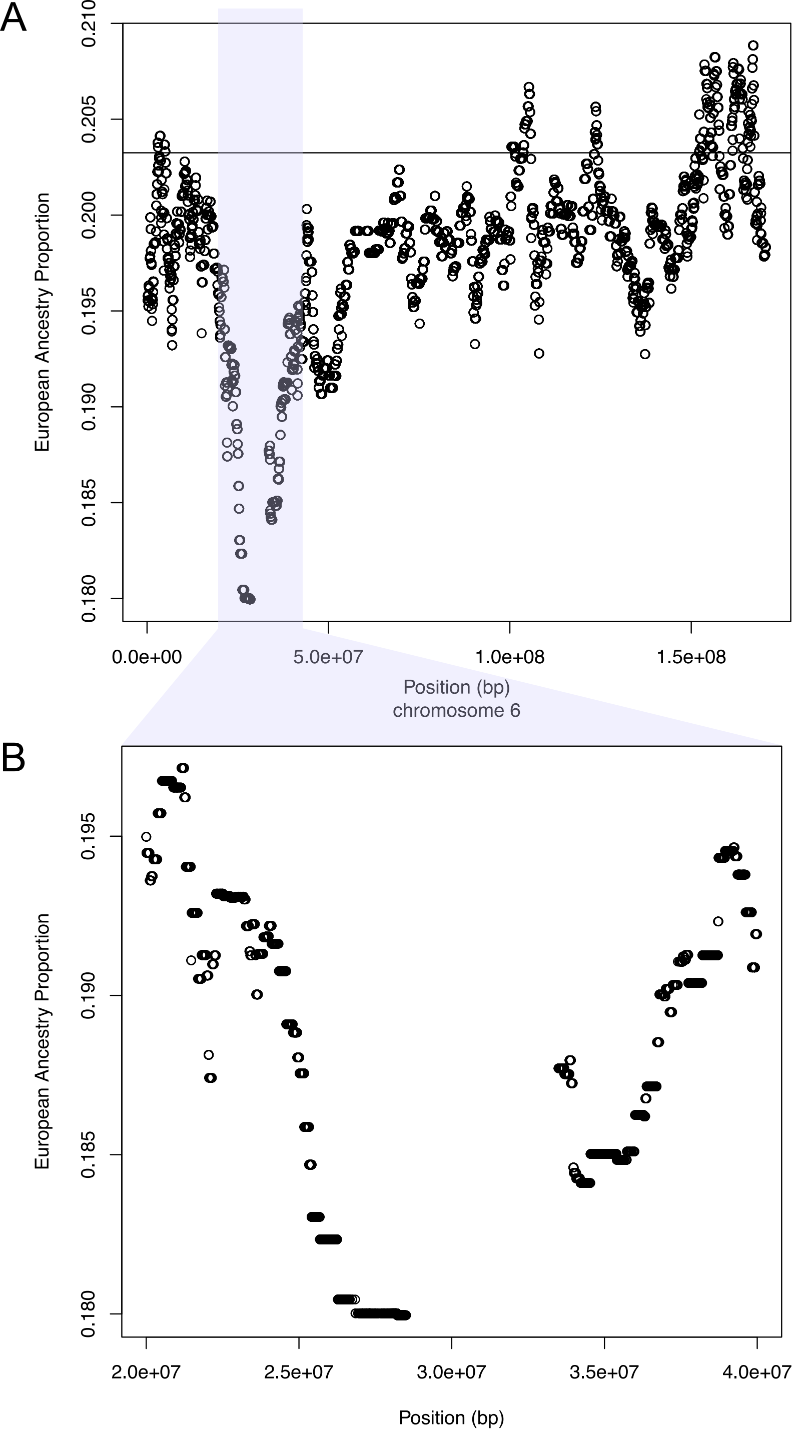

### Extended Data Fig. 5

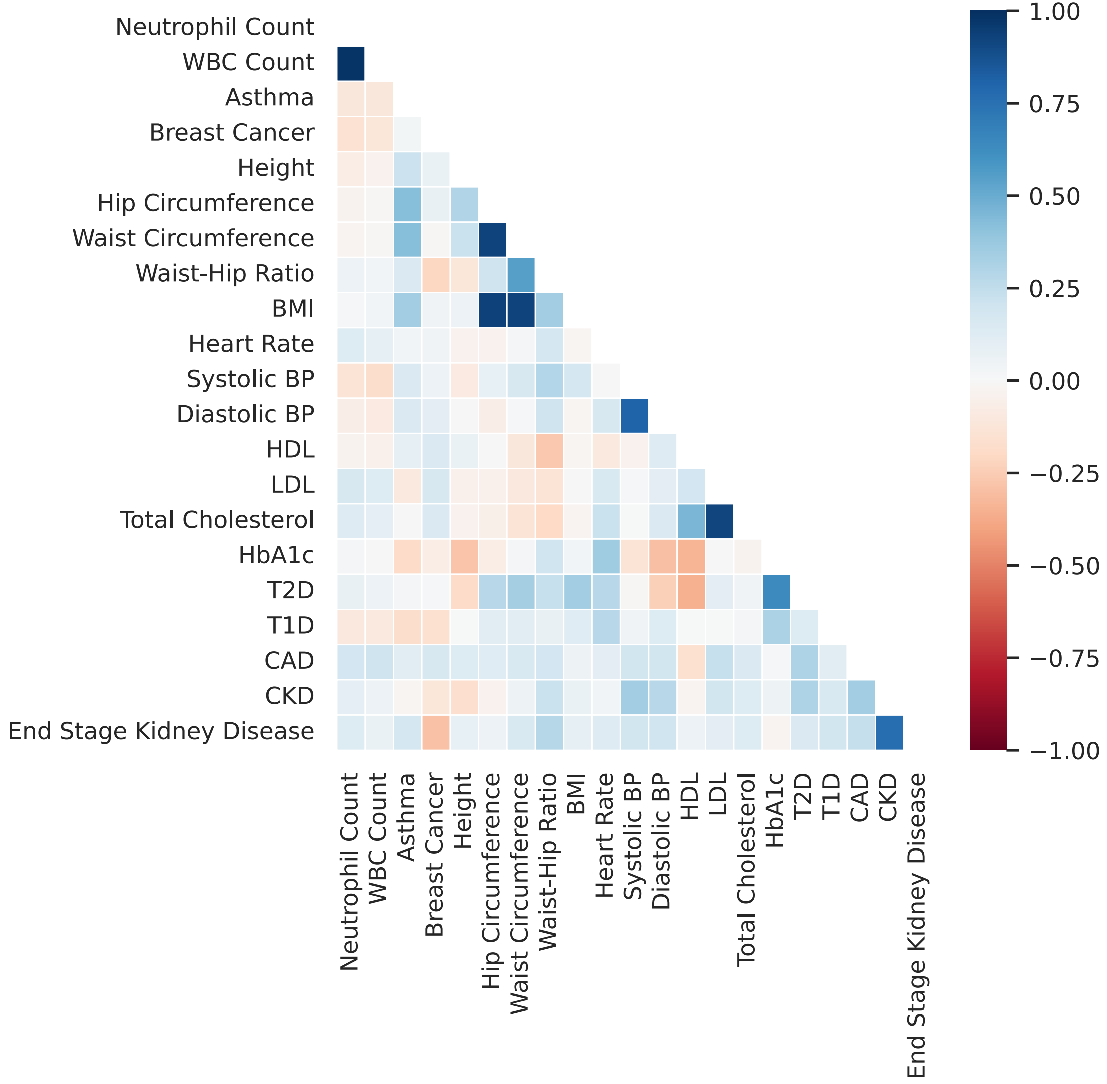

### Extended Data Fig. 6

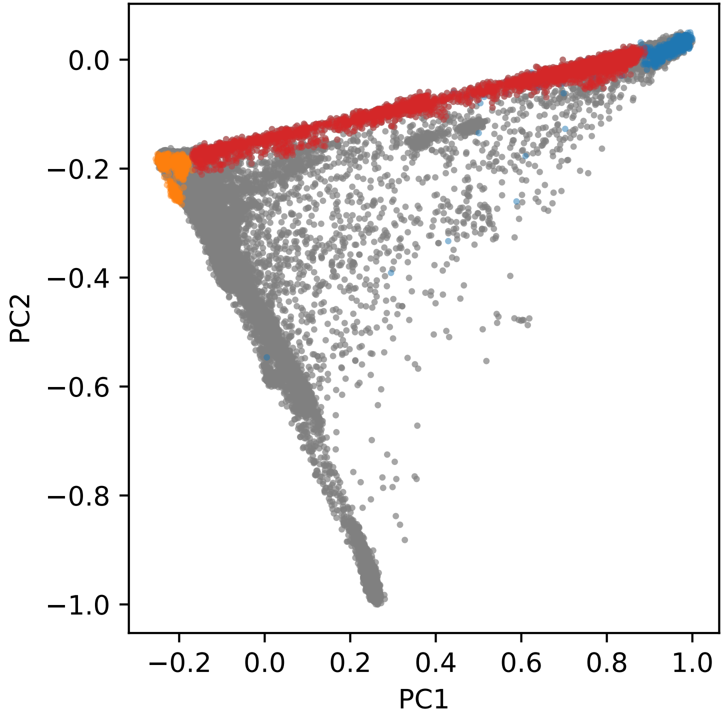

### Extended Data Fig. 7

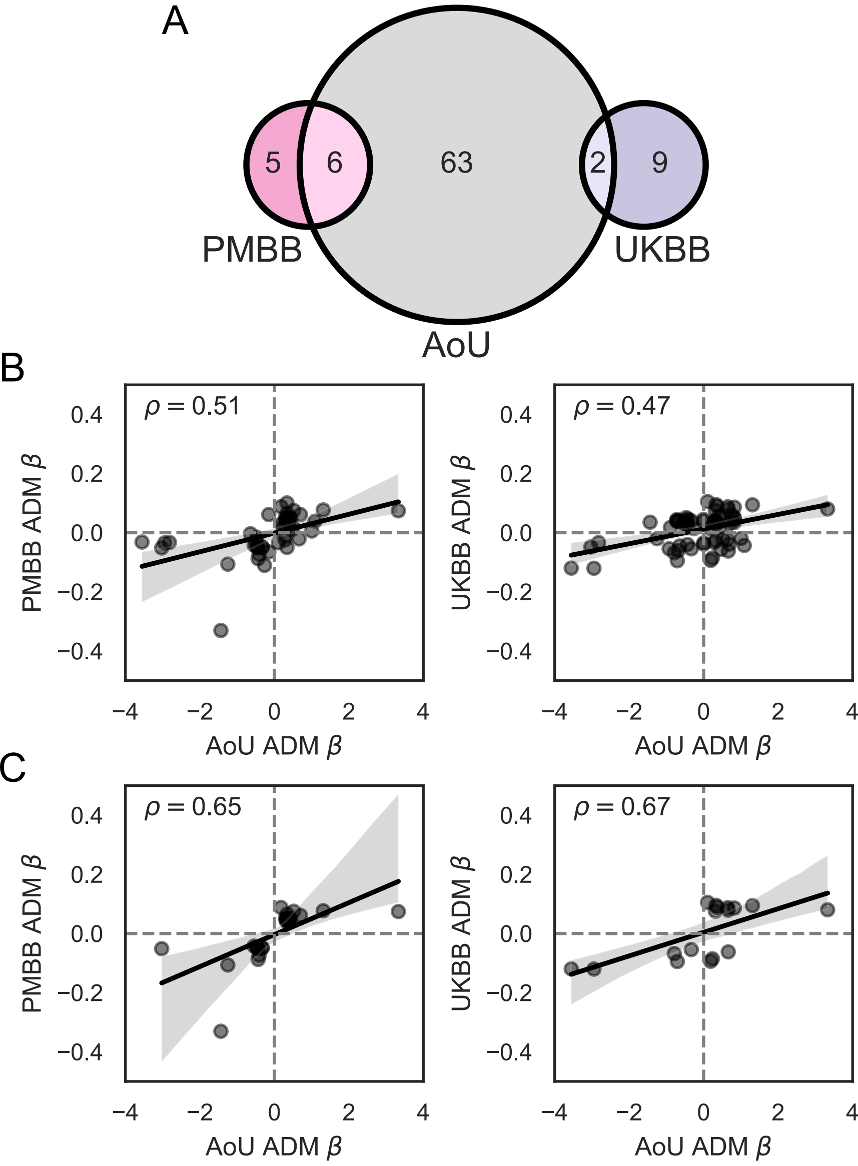

### Extended Data Fig. 8

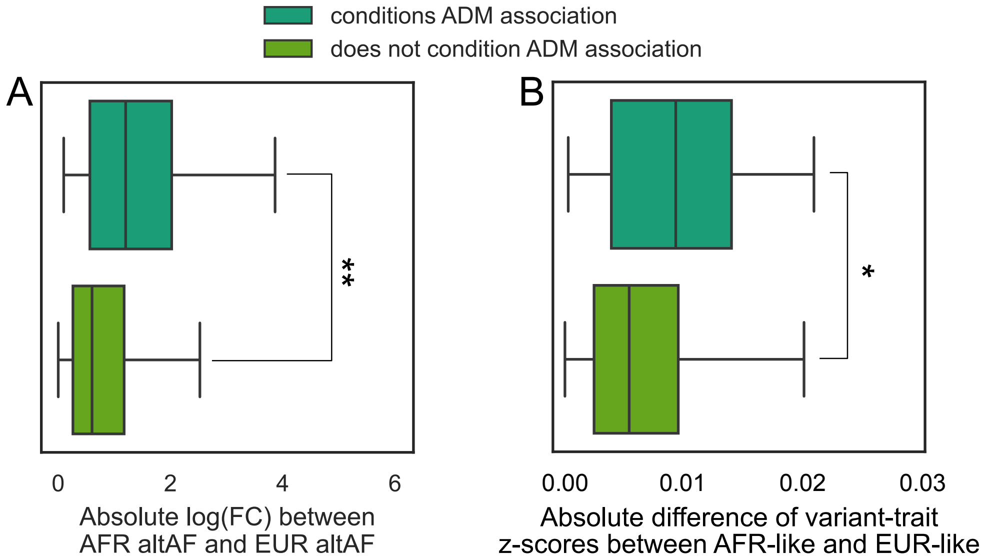
